# A benchmark of online COVID-19 symptom checkers

**DOI:** 10.1101/2020.05.22.20109777

**Authors:** Nicolas Munsch, Alistair Martin, Stefanie Gruarin, Jama Nateqi, Isselmou Abdarahmane, Rafael Weingartner-Ortner, Bernhard Knapp

## Abstract

**Background:** A large number of online COVID-19 symptom checkers and chatbots have been developed but anecdotal evidence suggests that their conclusions are highly variable. To our knowledge, no study has evaluated the accuracy of COVID-19 symptom checkers in a statistically rigorous manner.

**Methods:** In this paper, we evaluate 10 different COVID-19 symptom checkers screening 50 COVID-19 case reports alongside 410 non-COVID-19 control cases.

**Results:** We find that the number of correctly assessed cases varies considerably between different symptom checkers, with Symptoma (F_1_=0.92, MCC=0.85) showing the overall best performance followed by Infermedica (F_1_=0.80, MCC=0.61).

## Introduction

In the modern world, large numbers of patients initially turn to various online sources for self-diagnoses before seeking diagnoses from a trained medical professional. But web sources have inherent problems such as misinformation, misunderstandings, misleading advertisements and varying quality [1]. Interactive examples of web sources developed to meet the need of online diagnoses are sometimes referred to as symptom checkers or chatbots [2][3]. Based on a list of entered symptoms and other factors, symptom checkers return a list of potential diseases.

Online symptom checkers have become popular in the context of the novel coronavirus disease 2019 (COVID-19) pandemic as access to doctors is reduced, worry in the population is high, and lots of misinformation is circulating the web [1]. On COVID-19 symptom checker web pages users are asked a series of COVID-19 specific questions and, upon completion, an association between the answers and COVID-19 is given alongside behavioural recommendations, e.g., self-isolate.

COVID-19 symptom checkers are valuable tools for pre-assessment and screening during this pandemic, both taking pressure off from clinicians and reducing footfall within hospitals. A large number of symptom checkers specific to COVID-19 have been developed. Anecdotal evidence (e.g. a newspaper article [4]) suggests that their conclusions differ with possible implications on the quality of the symptom assessment. To our knowledge, there exist no studies comparing and evaluating COVID-19 symptom checkers.

In the following, we present a study evaluating 10 different COVID-19 online symptom checkers using 50 COVID-19 cases extracted from the literature and 410 non-COVID-19 control cases of patients with other diseases. We find that the COVID-19 symptom checkers’ classification of many patient cases differ as well as their accuracies. Symptoma (F1 =0.92, MCC=0.85) shows the overall best performance followed by Infermedica (F1=0.80, MCC=0.61).

## Methods

### COVID-19 symptom checkers

Ten COVID-19 symptom checkers that were freely available online between 3rd and 9th of April 2020 were selected for this study (Table 1) These symptom checkers were used in the versions available in this date range and updates after this date were not considered for analysis.

**Table 1:**
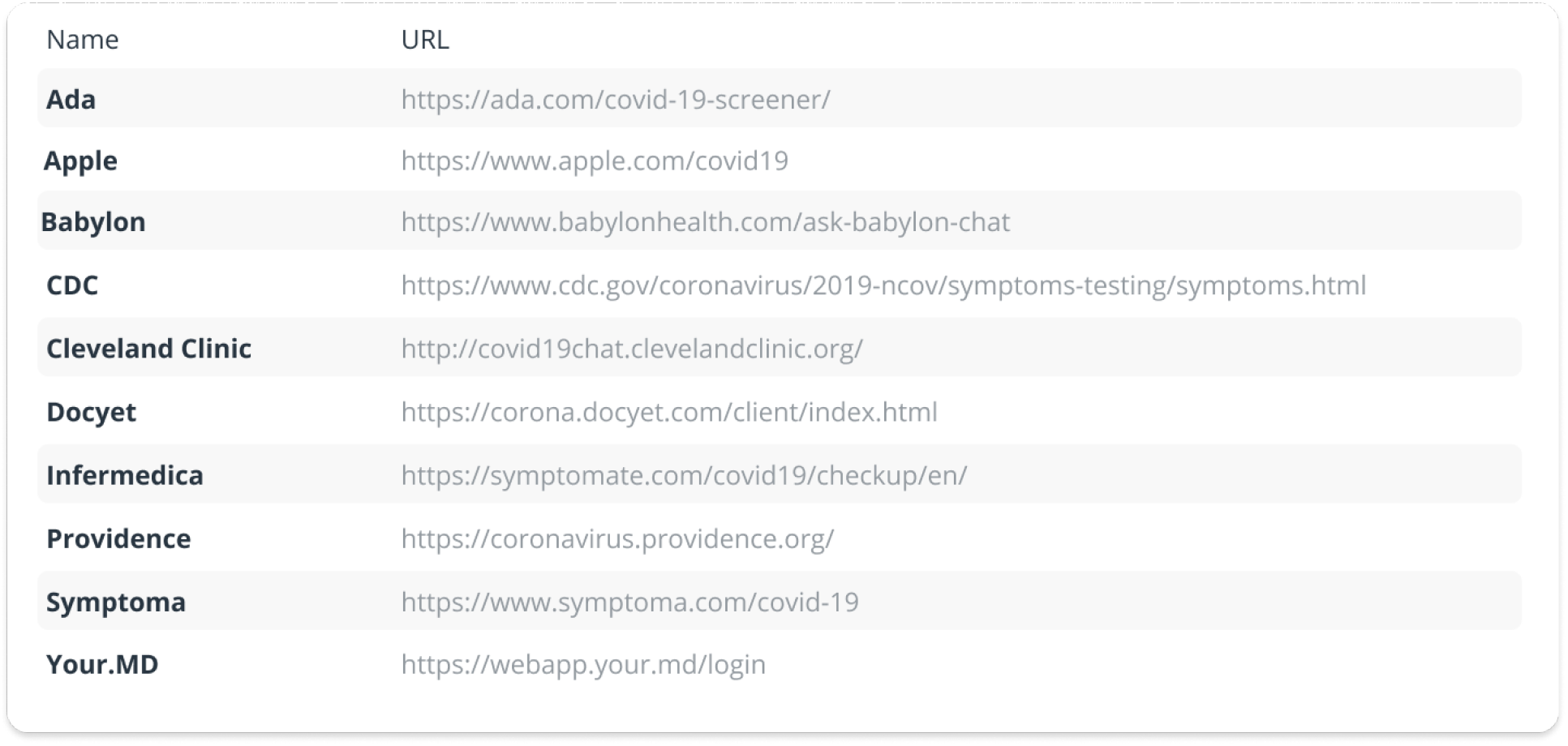
List of online COVID-19 symptom checkers included in this study.

As a baseline for the performance evaluation of the 10 online COVID-19 symptom checkers, we developed two additional simplistic symptom checkers. These two checkers evaluate and weigh the presence of WHO [5] provided COVID-19 symptom frequencies (see S1 Table) based on vector distance (SF-DIST) and cosine similarity (SF-COS). These approaches can be implemented in a few lines of code (see S2 Text).

### Clinical cases

We used a total of 460 clinical cases to evaluate the performance of the COVID-19 symptom checkers. Each case lists both symptoms and the correct diagnosis, alongside the age and sex of the patient when available. Details of the two case sets used are given below and in Table 2.

**Table 2:** Number of symptoms in case sets

| case set | Number of cases | Number of symptoms | Age | Sex |
| --- | --- | --- | --- | --- |
| **COVID-19 cases** | 50 | 8.2 ± 4.1<br>(median 7) | 45.6 ± 16.9<br>(median 45) | 50% male<br>42% female<br>8% unknown |
| **BMJ control cases** | 410 | 9.8 ± 4.4<br>(median 9) | 38.6 ± 22.4<br>(median 38) | 58% male<br>39% female<br>3% unknown |

### COVID-19 cases

A total of 50 COVID-19 cases were extracted by three trained medical doctors from the literature and are listed in S3 Table. Each case describes one patient’s medical situation, i.e. symptoms experienced or COVID-19 contacts. Extreme edge cases of COVID-19 such as patients with several severe comorbidities were not included in this study.

### Control cases

COVID-19 cases allow us to evaluate the sensitivity of symptom checkers. To also evaluate the specificity, 410 control cases from the British Medical Journal (BMJ) were sourced [6,7]. To allow a fair assessment, we only used cases containing at least one of the COVID-19 symptoms (see S4 Table) reported by the WHO [5]. Classifying non-relevant cases (e.g., a fracture) would overestimate the symptom checkers’ specificity. Furthermore, these patients would not consult an online COVID-19 symptom checker. None of these 410 BMJ cases has COVID-19 listed as the diagnosis as the cases where collected before the COVID-19 outbreak.

### Accuracy evaluation

For statistical analysis we used the following classification:

- True-positive (TP): COVID-19 case classified as COVID-19
- False-positive (FP): non-COVID-19 case classified as COVID-19
- True-negative (TN): non-COVID-19 case classified as non-COVID-19
- False-negative (FN): COVID-19 case classified as non-COVID-19

For each symptom checkers, we calculate the following metrics:

Sensitivity (= true positive rate) = 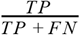

Specificity (= true negative rate) = 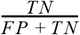

F1 score (= harmonic mean of the precision and recall) = 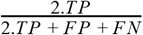

Matthews Correlation Coefficient (MCC) = 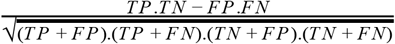

### Classification of symptom checkers’ outputs

Most COVID-19 symptom checkers return a human-readable text which contains an association between entered symptoms and COVID-19. We classified these associations into three different categories: high risk, medium risk and low risk. Examples of a high, medium and low risk classifications are “There is a high risk that COVID-19 is causing your symptoms”, “Your symptoms are worrisome and may be related to COVID-19” and “There’s nothing at present to suggest that you have coronavirus (COVID-19). Please practice physical/social distancing” respectively. Our full text-output to risk mapping for all symptom checkers and all text outputs is given in S5 Table.

Some symptom checkers only have two possible outputs: COVID-19 risk or no COVID-19 risk. In order to make symptom checkers with three and two risk levels comparable we performed two analysis versions: (a) medium risk and high risk is treated as COVID-19 positive (and low risk as COVID-19 as negative) and (b) high risk is treated as COVID-19 positive (and low risk and medium risk as COVID-19 negative).

### Bootstrapping

To evaluate the robustness of our statistical measures and account for the unbalanced dataset, we performed bootstrapping across our cases. A total of 3000 random samples consisting of 50 COVID-19 cases and 50 control cases were created by sampling with replacement from the original set of 50 COVID-19 cases and the 410 control cases.

## Results

### Sensitivity and specificity

In order to analyse the performance of the 10 online symptom checkers, we calculated the sensitivity and the specificity of each symptom checker based on the cases described in the method section. A scatterplot between sensitivity and specificity to COVID-19 of the different symptom checkers is given in Fig 1 and detailed numerics in S6 Table and S7 Table. These symptom checkers fall roughly into four groups: upper left corner, lower right corner, central region, upper right corner.

**Fig 1.**
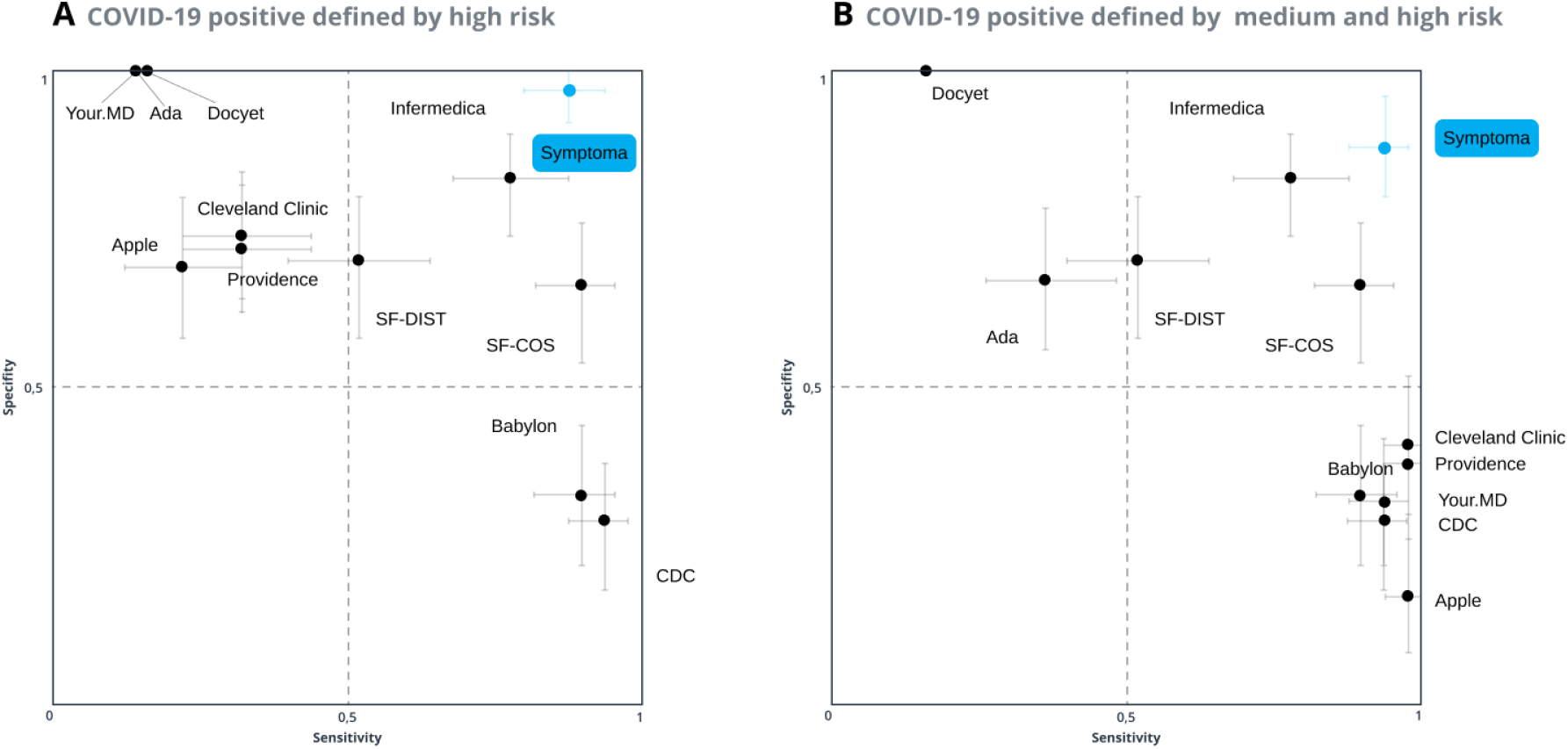
Sensitivity and specificity to COVID-19 cases. The mean of the 3000 random samples and 90% bootstrap confidence interval are reported as dots and crosses respectively. (A) High risk: A COVID-19 positive prediction is defined only by a high risk returned by a symptom checker. (B) Medium-High risk: A COVID-19 positive prediction is defined by either a medium risk or high risk returned by a symptom checker.

Further analysis of true and false case classifications of these groups shows that the group in the upper left corner is composed of symptom checkers that require one (or few) highly specific symptoms to be present in order to classify a case as COVID-19 positive (e.g. “intensive contact with a COVID-19 positive person”). By this way, these symptom checkers miss many COVID-19 positive patients that did not report exactly this highly specific symptom. Vice versa such highly specific symptoms are hardly present in non-COVID-19 cases. This results in low sensitivity and high specificity.

The group in the lower right corner is composed of symptom checkers which predict a case as COVID-19 positive based on the presence of one or few COVID-19 associated symptoms, e.g. the presence of fever or cough is enough to predict a patient as COVID-19 positive. These checkers classify nearly every patient that has a respiratory disorder or viral infection as COVID-19 positive. As such, they do not miss many COVID-19 patients but wrongly predict many non-COVID-19 patients as COVID-19 positive. This results in low specificity and high sensitivity.

The group in the more central region is composed of symptom checkers which use a more balanced prediction but exhibit limited success correctly classifying COVID-19 and non-COVID-19 patients.

The group in the upper right corner is composed of symptom checkers which also use a more balanced “symptoms to COVID-19 association model” but in this case, the classification between COVID-19 and non-COVID-19 patients is more successful.

### Constraining symptoms for Symptoma

As Symptoma exhibits the best combination of sensitivity and specificity, we focused our analysis on Symptoma’s performance. Symptoma allows free-text input of one’s symptoms and thereby a more precise representation of the clinical test cases. The other symptom checkers do not allow free text input which limits the number of possible symptoms considerably (Fig 2A). In order to investigate how Symptoma would perform if constrained, we performed pairwise comparisons where Symptoma is only allowed to use the symptoms of another symptom checker. In this setup, Symptoma is massively disadvantaged as it can not use its full abilities. For example, in the pairwise comparison with “Your.MD”, Symptoma considers only “fever”, “dry cough”, “shortness of breath”, and “contact with a confirmed COVID-19 case” for the classification of cases. The results of this analysis are summarised in Fig 2B. the sensitivity and specificity scatter plots are provided in the S8 Fig and detailed numerics in S9 Table and S10 Table. Under these constraints and when COVID-19 positive is defined by high risk only, Symptoma still significantly outperforms Apple and Cleveland Clinic, while performing statistically similar to six of the remaining symptom checkers (upper panel of Fig 2B) When COVID-19-positive is defined by high and medium risk (lower panel of Fig 2B) Symptoma’s constrained performance is similar to seven of the other checkers, while outperforming Ada and Docyet. For Apple, Babylon, CDC, Cleveland Clinic, Providence and “Your.MD” the performance is about the same. When Symptoma is allowed to use all symptoms of the case descriptions, it clearly outperforms all other checkers (dashed blue line in Fig 2B) This suggests that performance is directly related to the number of symptom’s any given checker considers as input, and as such, free-text input (non-constrained) will normally lead to a higher likelihood of correct diagnosis.

**Fig 2.**
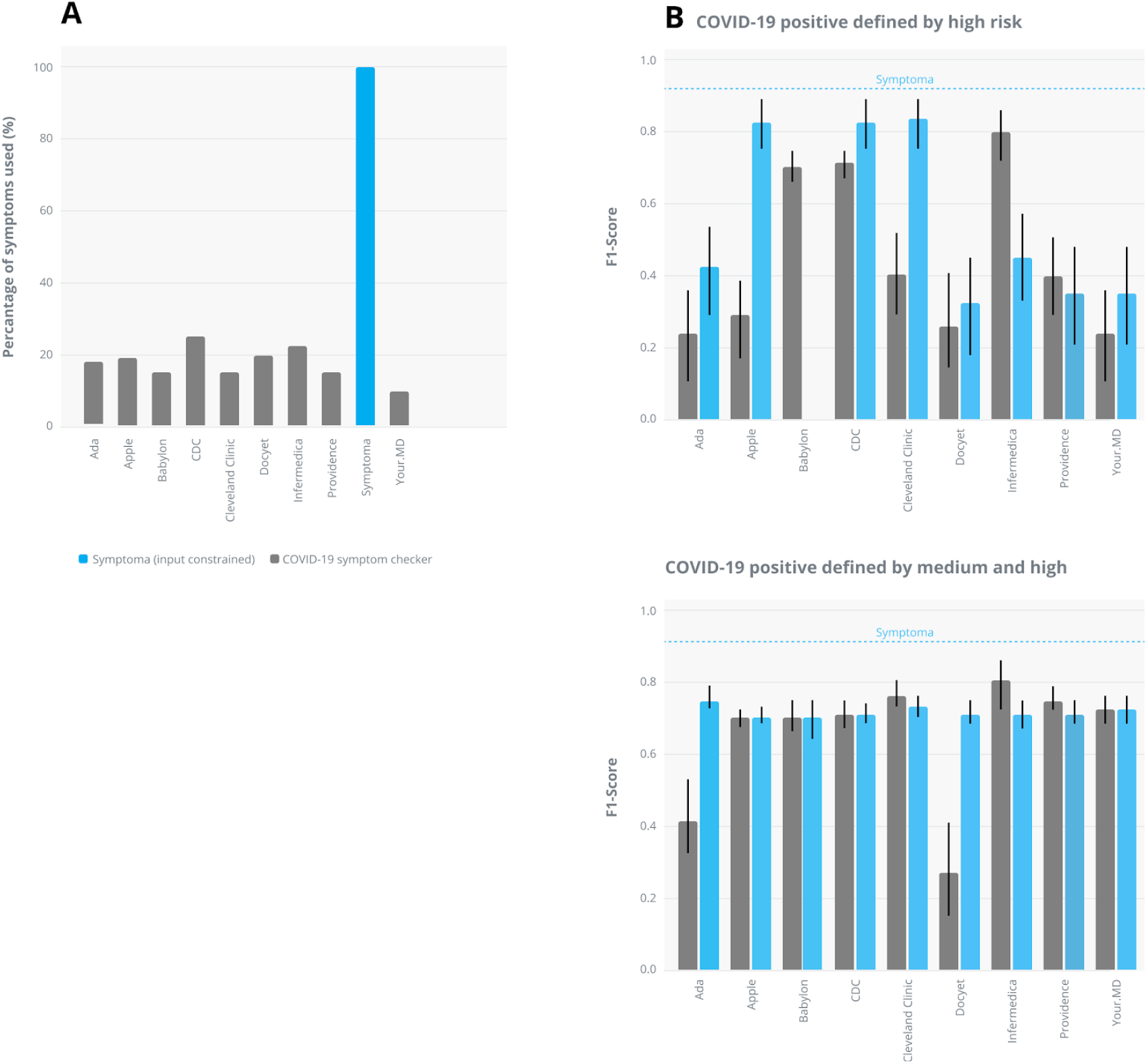
(A) Percentage of symptoms used for case classifications by each symptom checker relative to the total number of symptoms contained in all cases. (B) Symptoma input-constrained evaluation: Pairwise comparison between all symptom checkers and Symptoma based on the F1 score if only the subset of symptoms used by one checker is also used for Symptoma. The same analysis based on the MCC is shown in the S11 Fig. Please note that using only Babylon’s symptom inputs all cases are either classified medium or low risk by Symptoma and therefore there is no bar in the upper panel for Babylon’s Symptoma.

## Discussion

We classified 50 COVID-19 case descriptions from the recent literature as well as 410 non-COVID-19 control cases with ten different online COVID-19 symptom checkers. Only two out of ten symptom checkers showed a reasonably good balance between sensitivity and specificity: namely Infermedica (F_1_=0.80) and Symptoma (F_1_=0.92). Most other checkers are either too sensitive, classifying almost all patients as COVID-19 positive, or too specific, classifying many COVID-19 patients as COVID-19 negative (see Fig 1). For example, our BMJ control cases contain a patient suffering from a pulmonary disease who presents with various symptoms, including fever, cough and shortness of breath, the three most frequent symptoms associated with COVID-19. Symptoma uses the additional symptoms and risk factors not considered by the other checkers, namely loss of appetite, green sputum, and a history of smoking, to discern the correct diagnosis of COVID-19 negative. Five of the other checkers consider this case as high risk.

Furthermore, most of the symptom checkers are even out-performed by our simplistic symptom frequency vector approaches (SF-DIST (F_1_=0.57) and SF-COS (F_1_=0.79)). Notably, the cosine version shows surprisingly good results outperforming 8 out of 10 symptom checkers based on the F_1_, score.

To our knowledge this is the first scientific evaluation of online COVID-19 symptom checkers, however, there are a number of related studies evaluating symptom checkers. These include a study that evaluated 23 general-purpose symptom checkers based on 45 clinical case descriptions across a wide range of medical conditions and found that the correct diagnosis was on average listed among the top 20 results of the checkers in 58% of all cases [2]. This study design was extended for five additional symptom checkers using ear, nose and throat (ENT) cases showing similar results [8]. Other evaluations include symptom checkers used for knee pain cases that found, based on 527 patients and 26 knee problems, that the physician’s diagnosis was present within the prediction list in 89% of the cases while the specificity was only 27% [9]. In another study, an analysis of a university students’ automated self-assessment triage system prior to an in-person consultation with a medical doctor found that the system’s urgency rating agreed perfectly in only 39% of cases while for the remaining cases the system tended to be more risk averse than the doctor [10]. Also, the applicability of online symptom checkers for 79 persons aged ≥50 years based on “think-aloud” protocols [11], deep learning algorithms for medical imaging [12], and services for urgent care [3] were evaluated.

If the performance of any (COVID-19) online symptom checker is acceptable depends on the perspective and use of the results. In the case of COVID-19, an online assessment can not fully replace a PCR-test as some people are asymptomatic, while others presenting with very specific COVID-19 symptoms might, in fact, have a very similar but different disease. Regardless, online COVID-19 symptom checkers can act as a first triage shield to take pressure off from in-person physician visits or hospitals. Symptom checkers could even replace telephone triage lines in which non-medically trained personnel read a predefined sequence of questions. Even though this was not part of this study, the authors believe that COVID-19 symptom checkers (if appropriately maintained and tested) might also be more reliable than the direct use of search engines such as Google or information via social media.

The strength of this study lies in the fact that it is based on a large number (n=460) of real patients’ case descriptions from the literature and a detailed evaluation on the best performing symptom checker (Fig 2). Vice versa, a potential weakness of this study lies in using real literature-based cases, which might have biased the test set to rather severe cases of COVID-19, as mild and uninteresting cases are usually not found in the literature. We countered this bias by not including extreme edge cases from the literature into our 50 COVID-19 cases. Another bias might be that our control case descriptions do not report a “COVID-19 contact”, even though a person with, for example a cold, might have had a COVID-19 contact (and did not get infected). Another limit of this study is the non-straight forward mapping of the symptom checker outputs to risk levels (S5 Table). The interpretation of the textual output is debatable in some cases. We countered this by allowing three different risk levels and merging them together in two different ways (see Fig 1A and Fig 1 B). We also classified every symptom checker output by multiple persons until consensus was reached.

## Conclusion

Symptom checkers are being widely used in response to the COVID-19 global pandemic. As such, quality assessment of these tools is critical. We show that various online COVID-19 symptom checkers vary widely in their predictive capabilities, with some performing equivalently to randomly guessing, while others, namely Symptoma (F_1_ = 0.92) and Infermedica (F_1_ = 0.80), exhibiting high accuracy.

## Data Availability

All analysis can be reproduced via the web-interfaces given in Table 1

## Contributors

Study design: BK, AM, JN, NM. Data compilation: SG, NM, RW, IA. Data analysis: NM, AM, BK, IA, RW. Writing the manuscript: BK, NM. Revising the manuscript critically: AM, BK, SG, JN.

## Acknowledgments

This study has received funding from the European Union’s Horizon 2020 research and innovation programme under grant agreement No 830017.

## Supporting information

**S1 Table.**
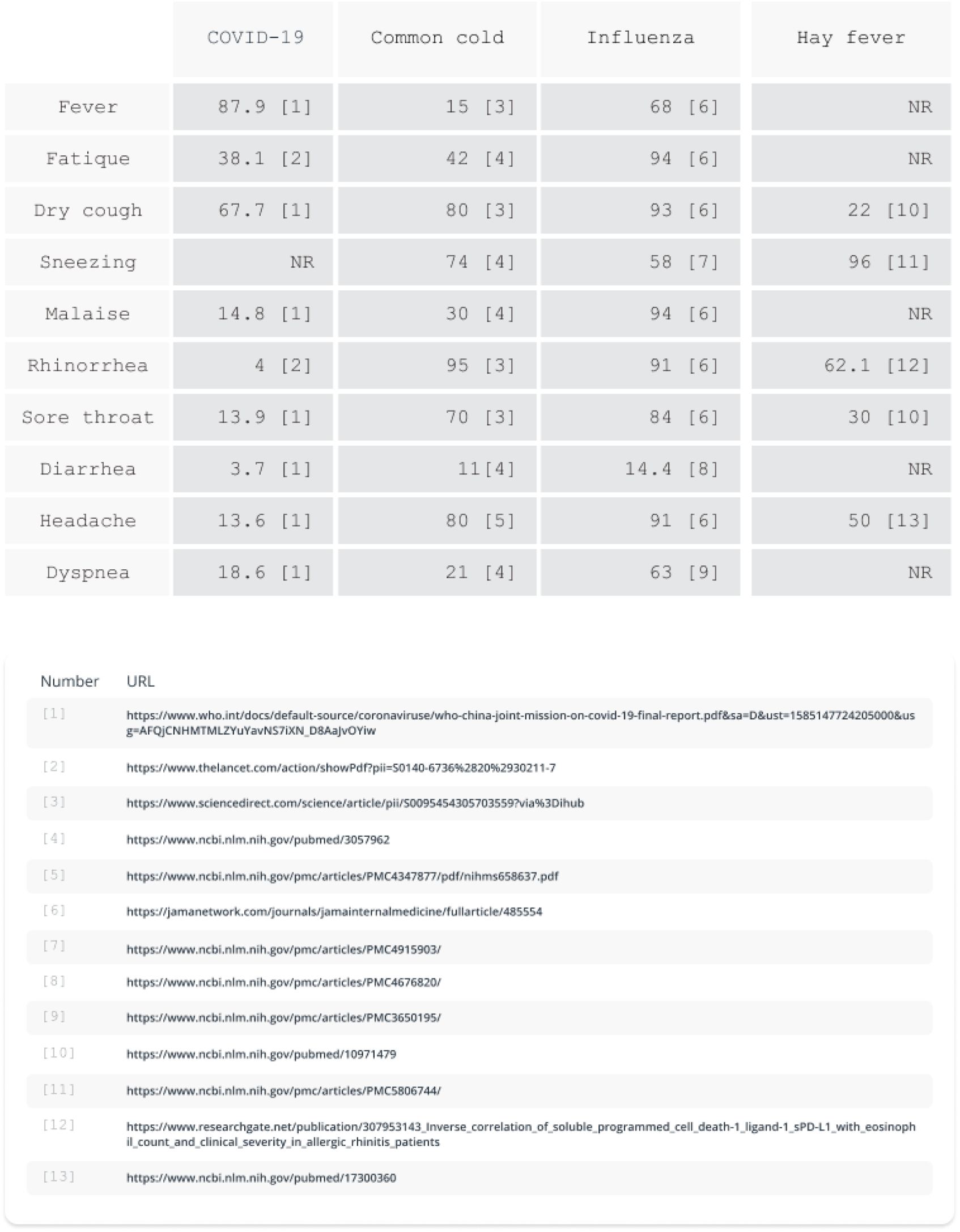
Symptom frequencies used in S2 Text.

**S2 Text.**
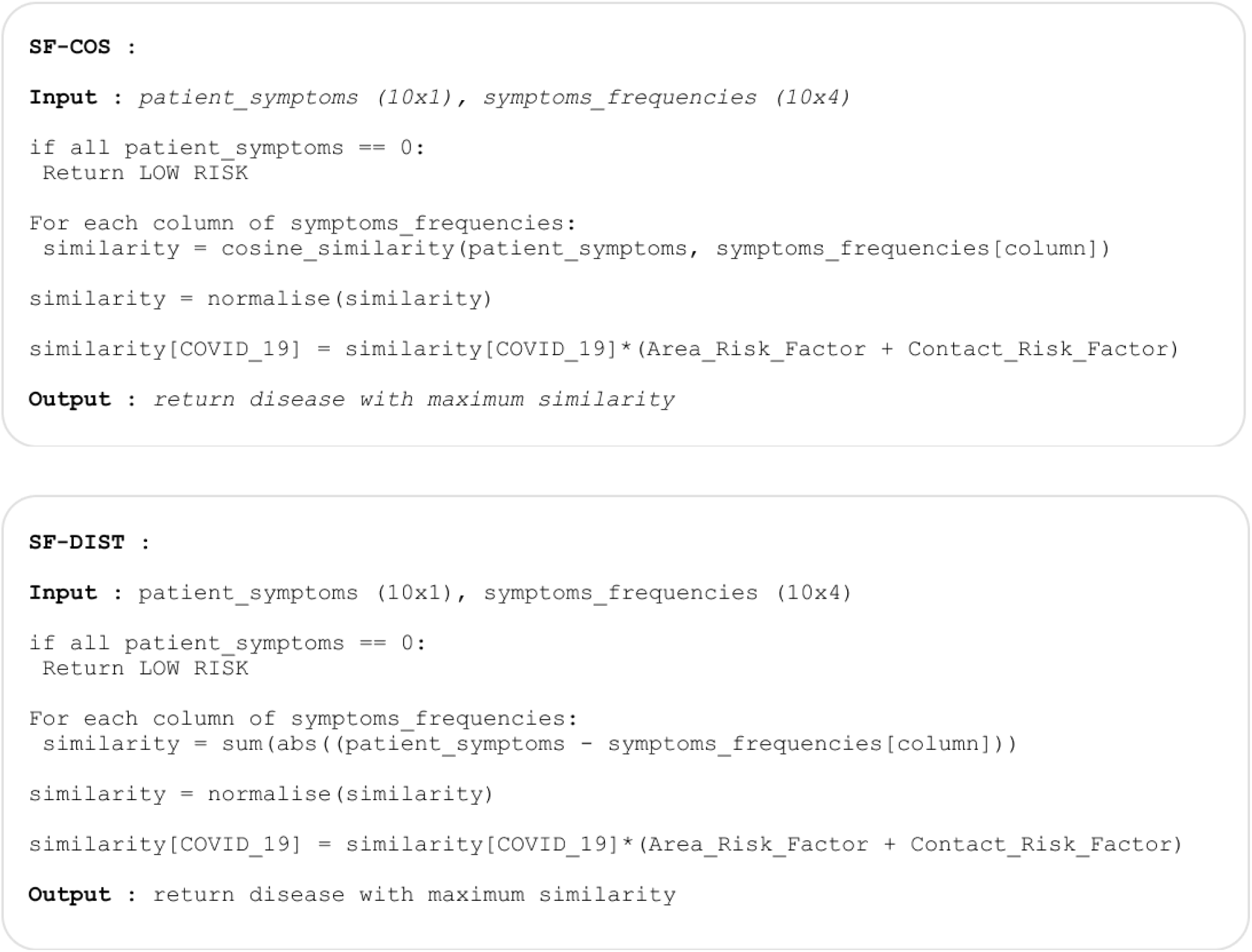
Pseudo code of symptom frequencies based on vector distance (SF-DIST) and cosine similarity (SF-COS)

**S3 Table.**
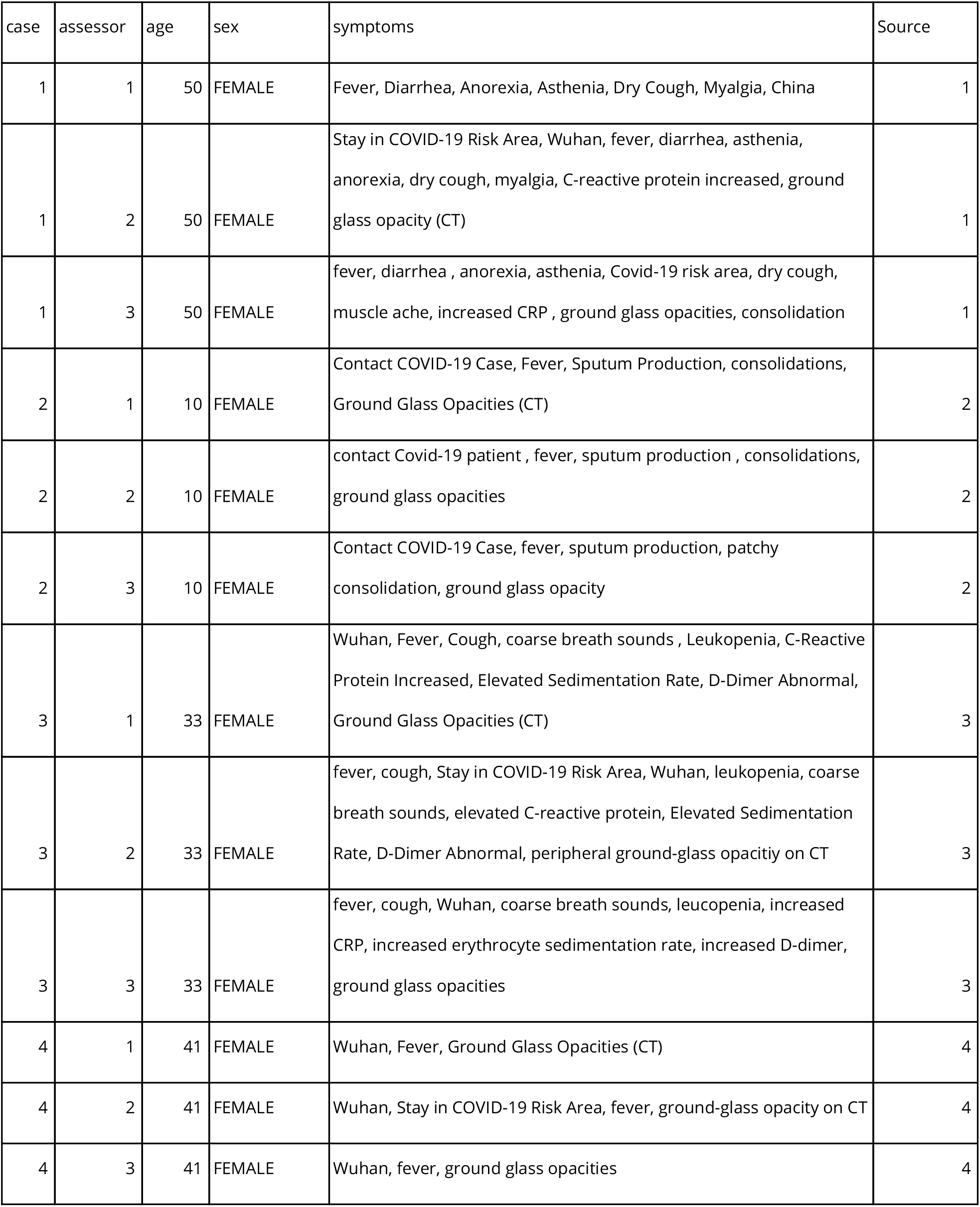
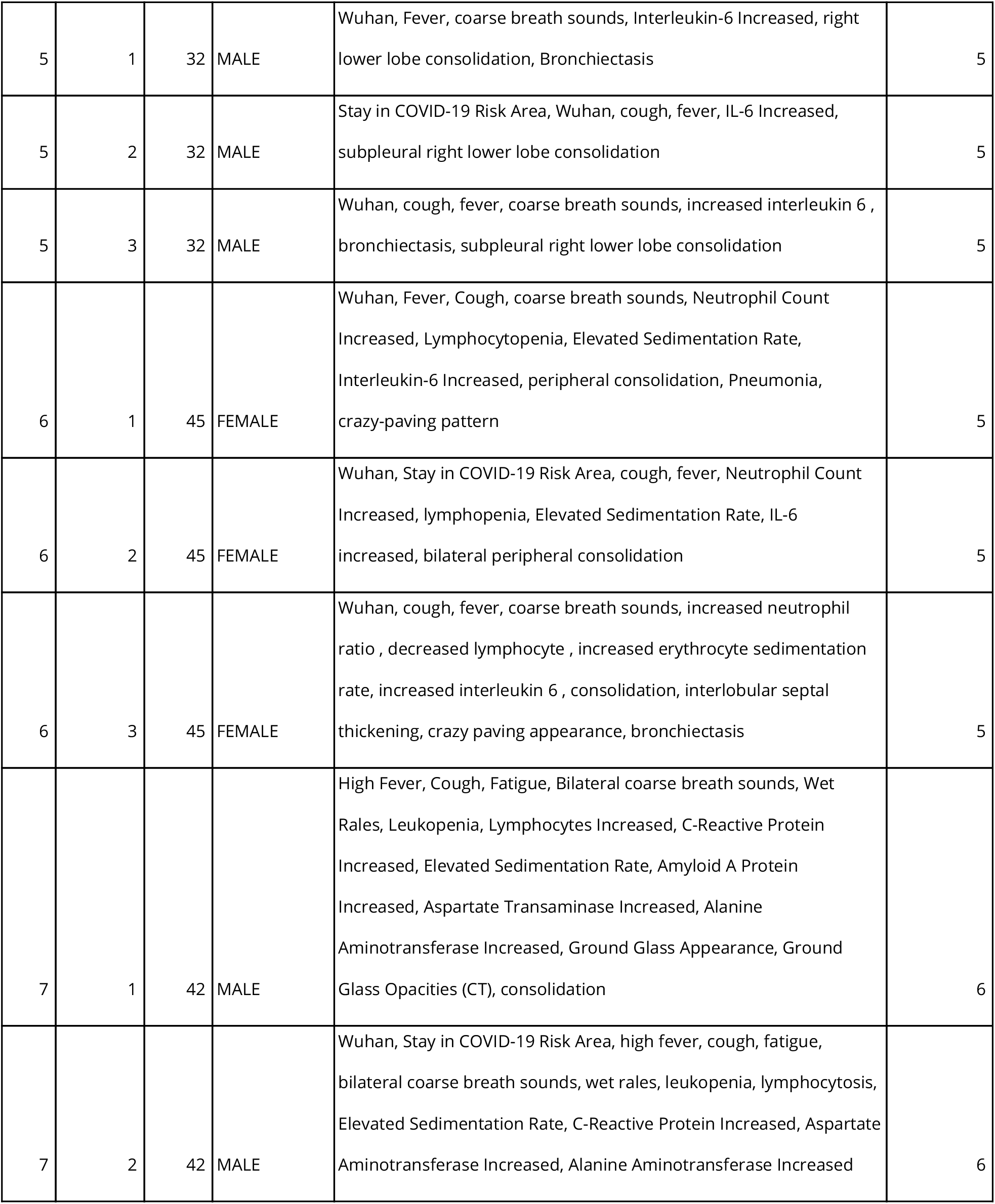
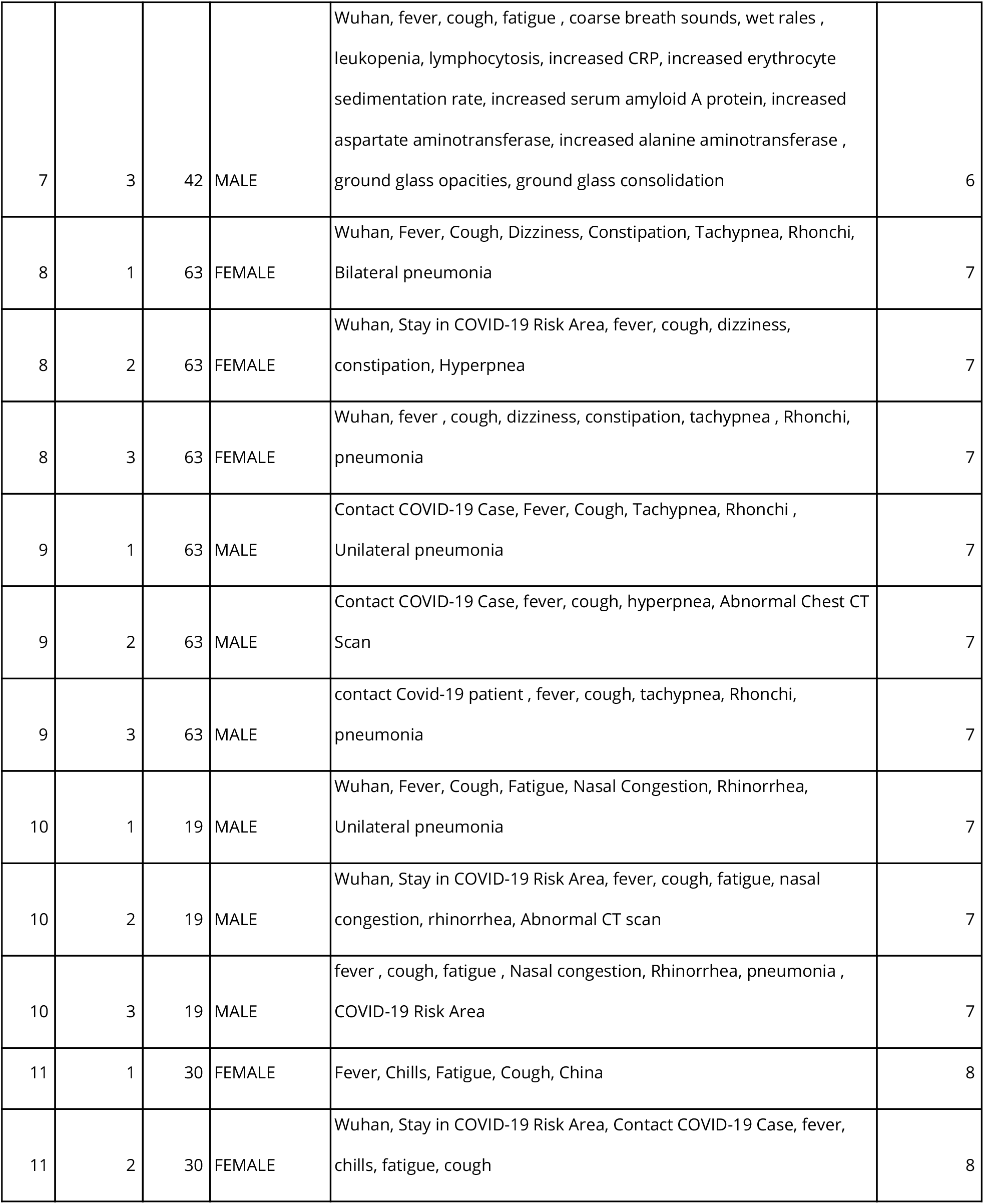
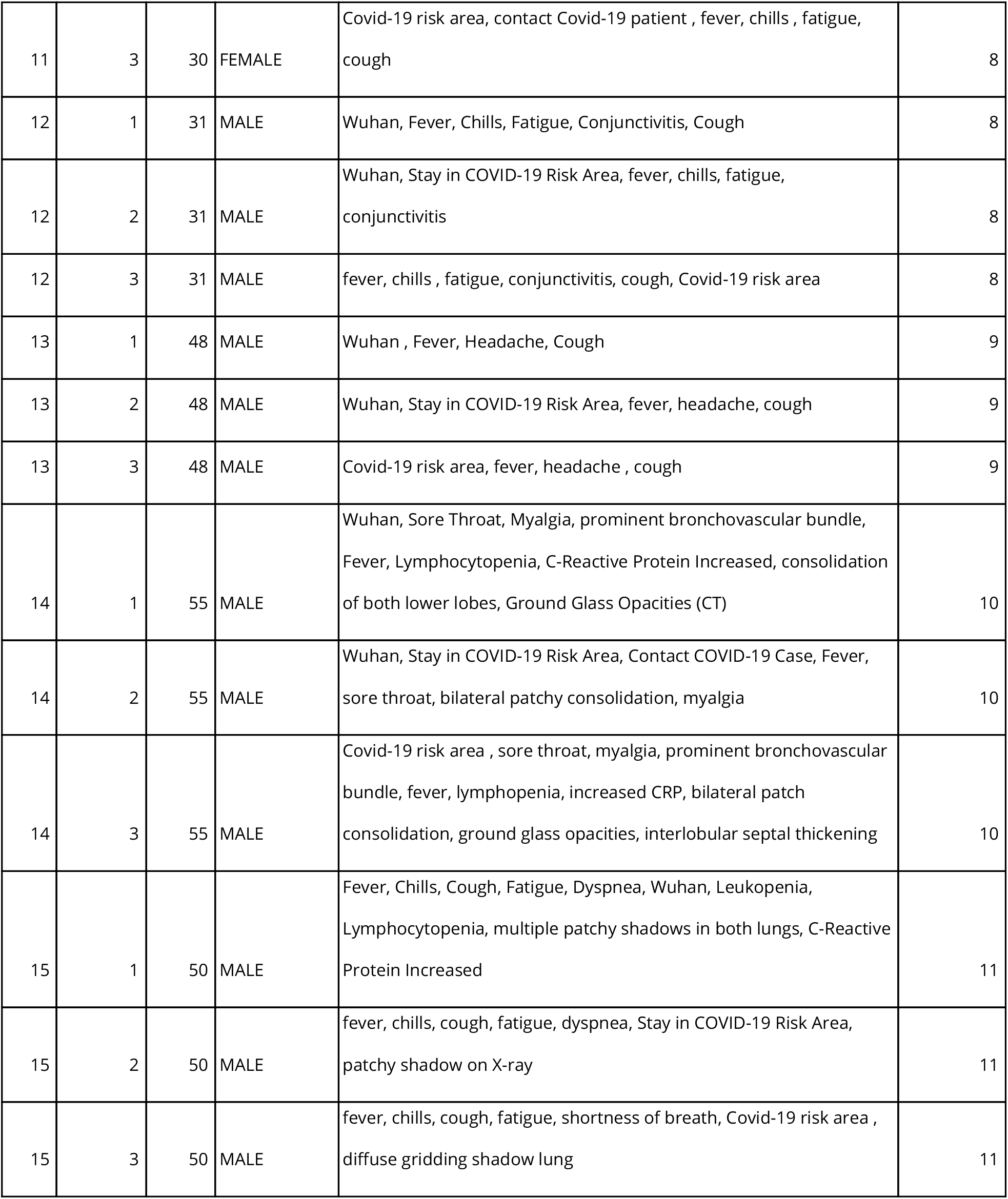
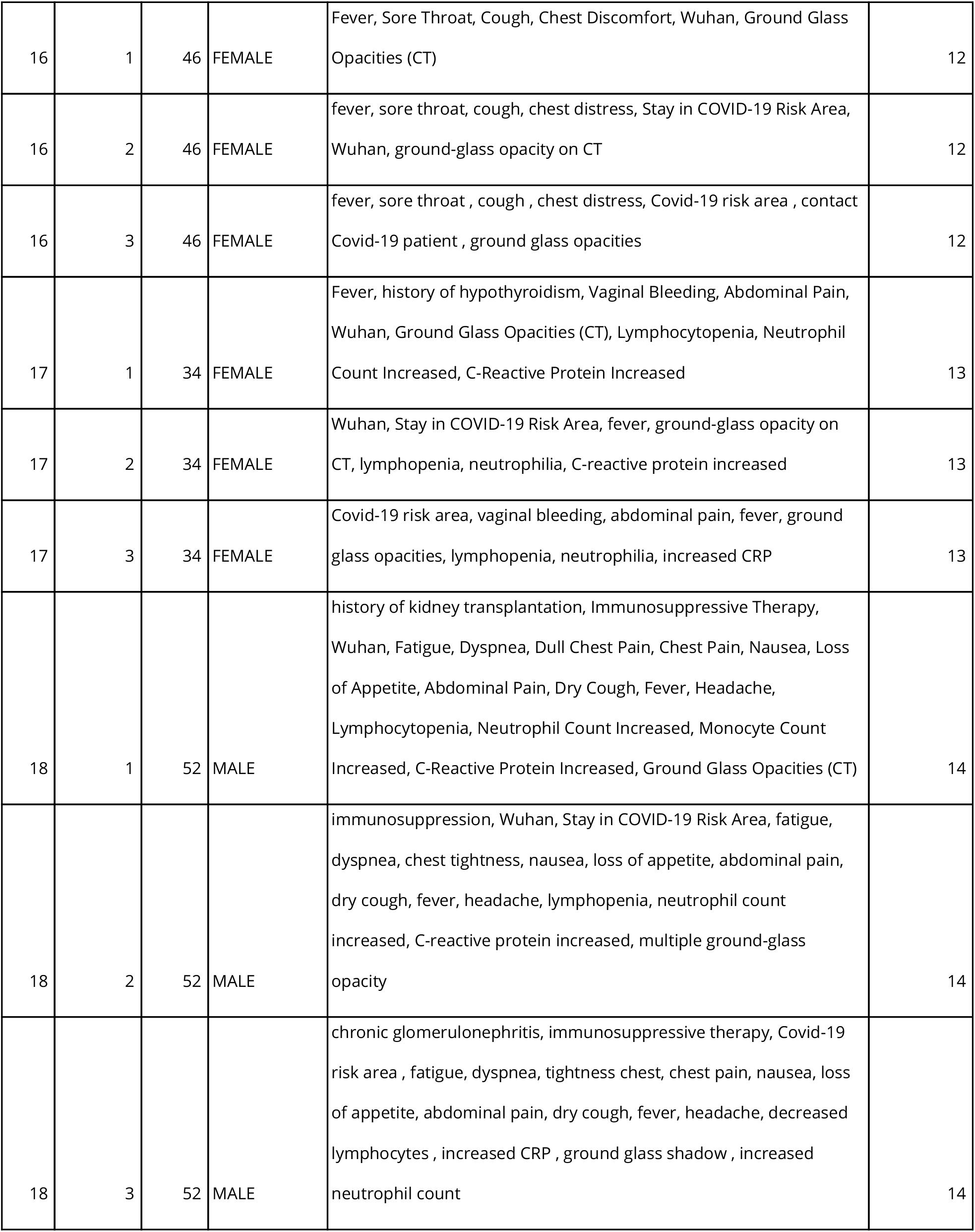
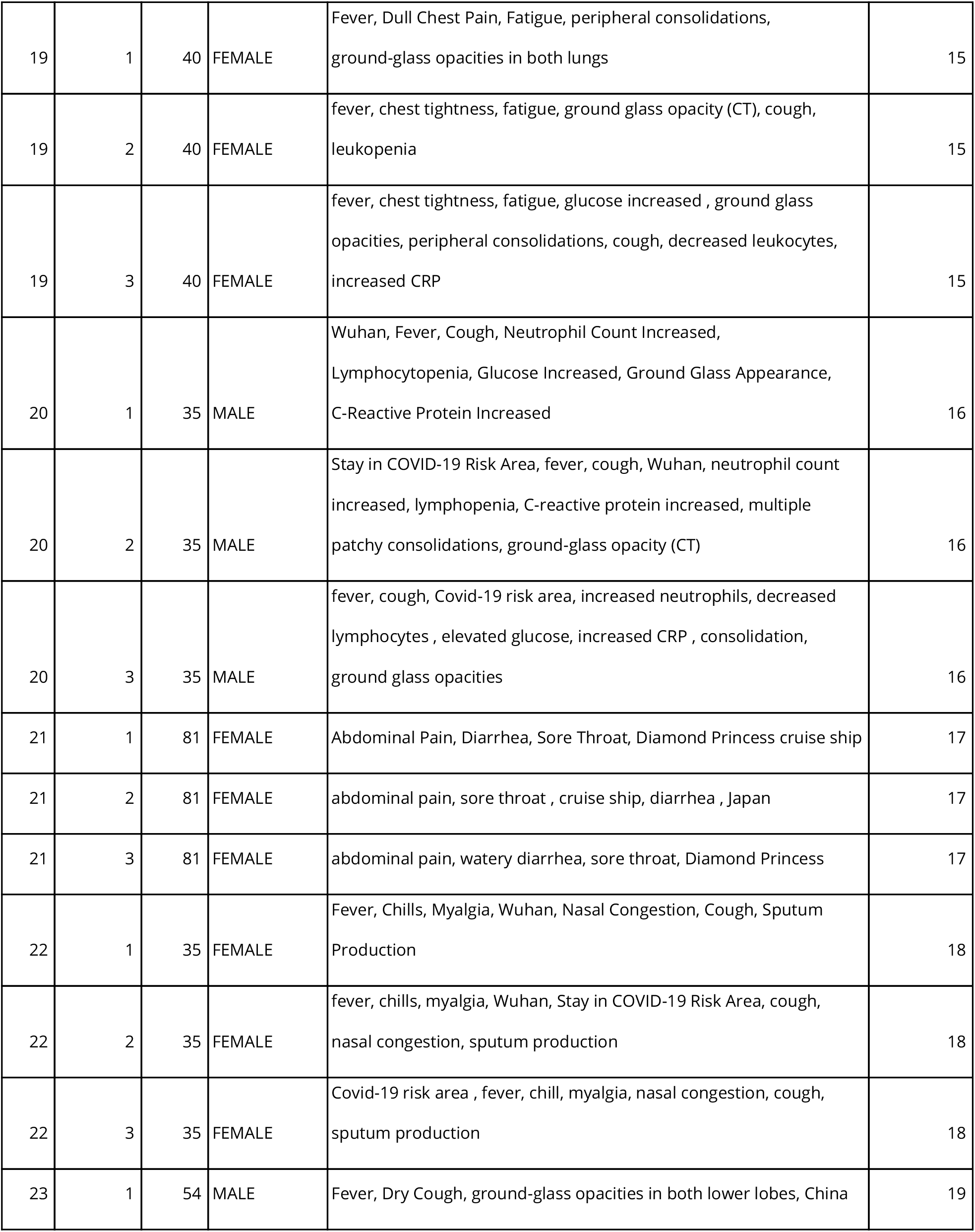
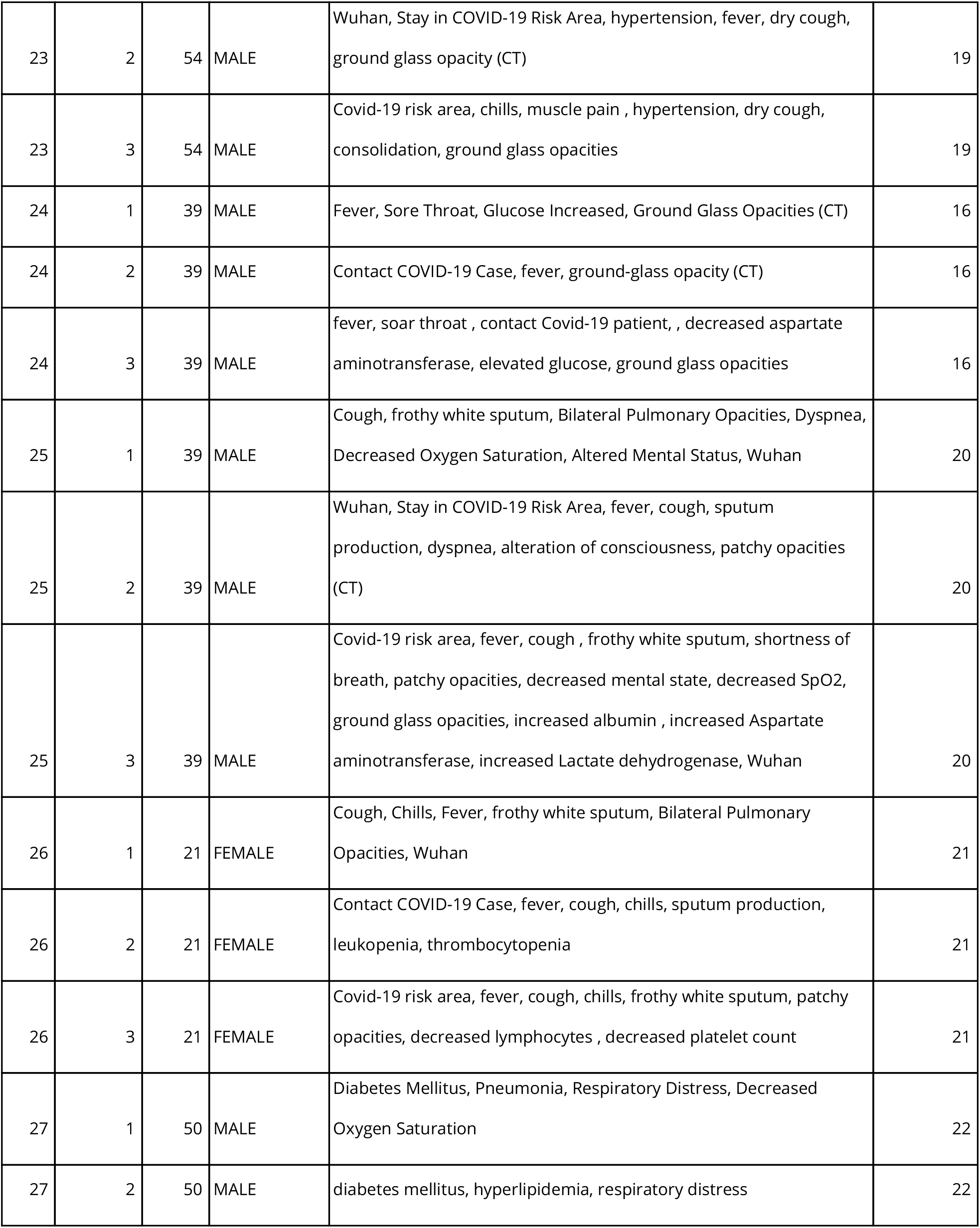
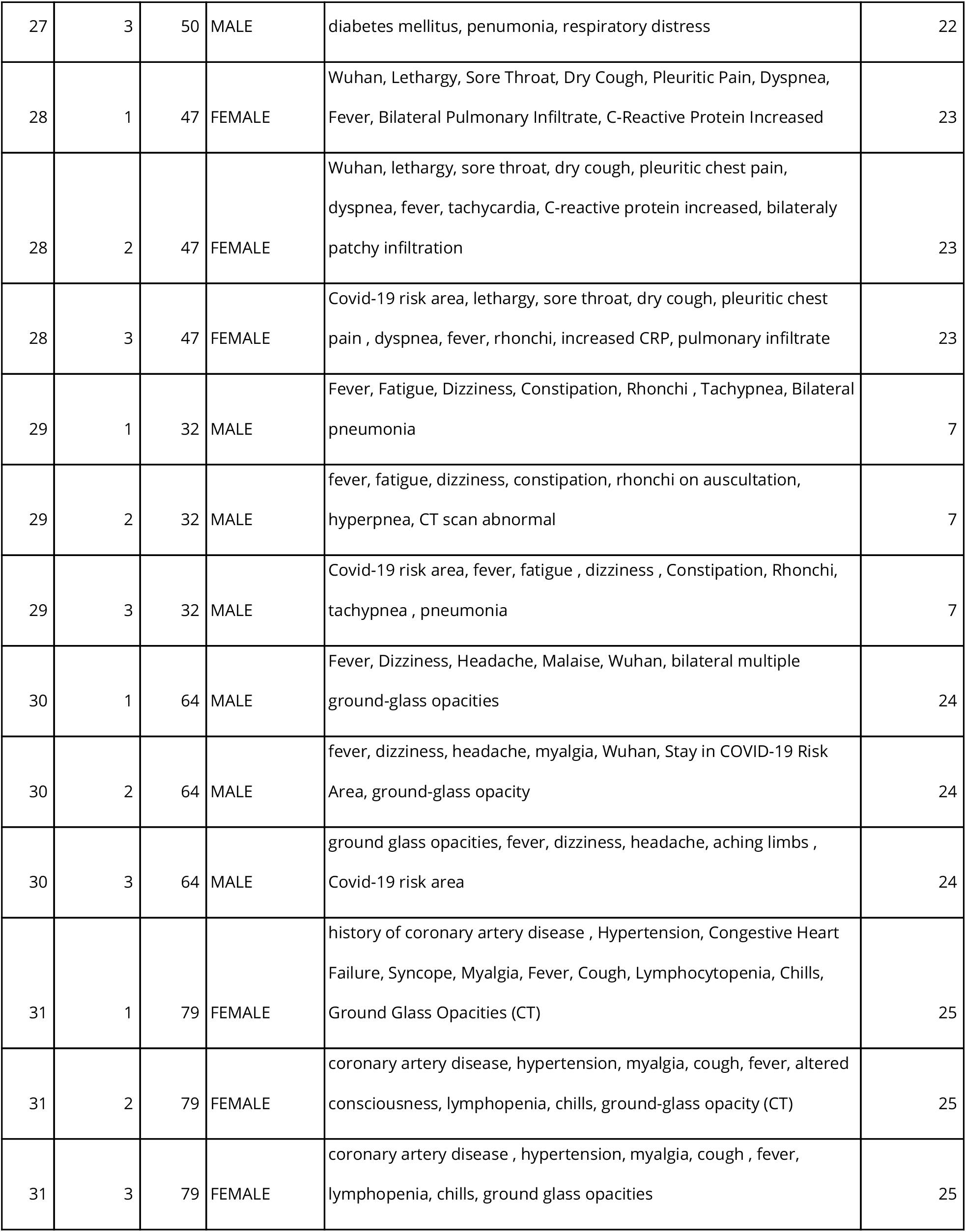
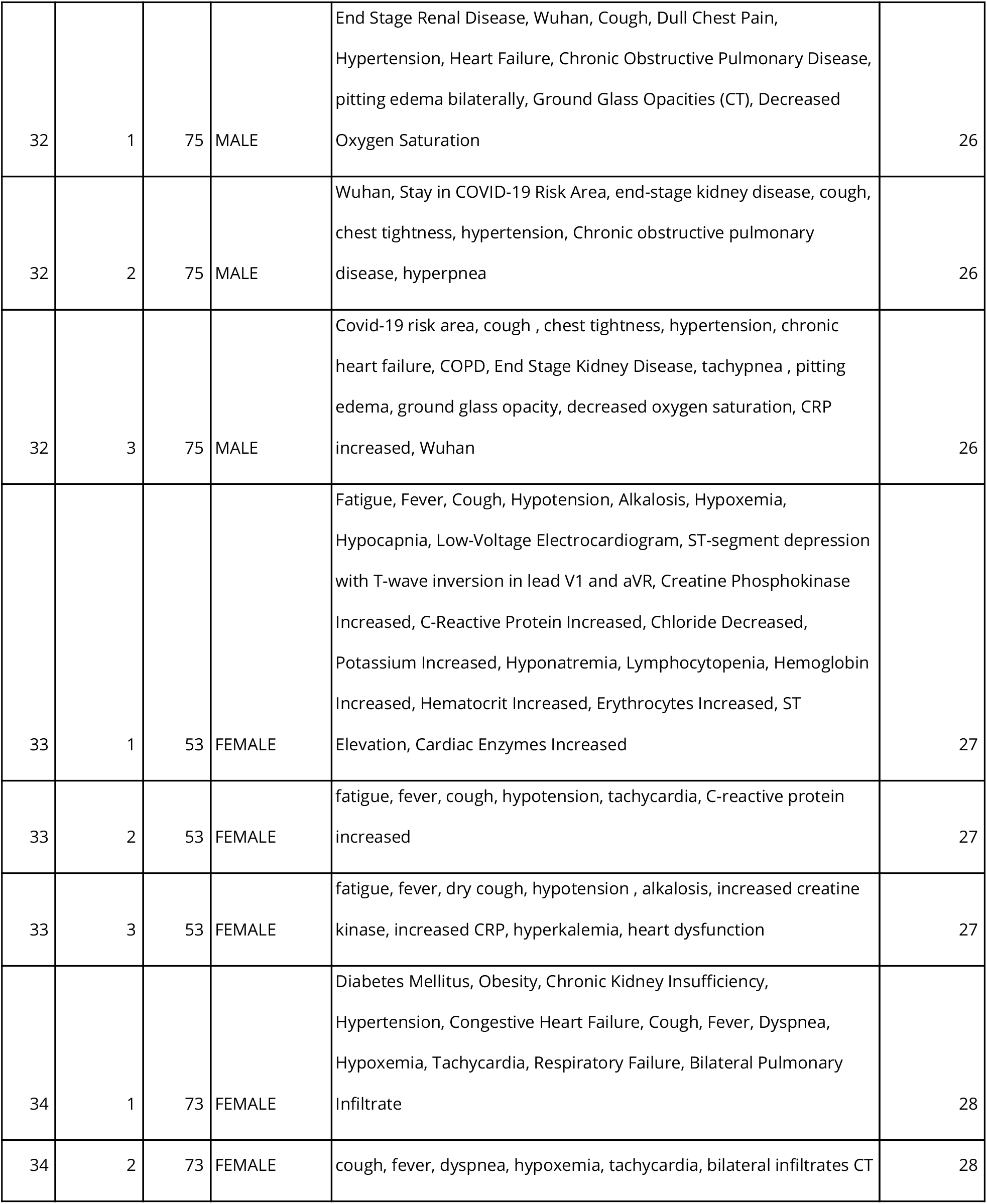
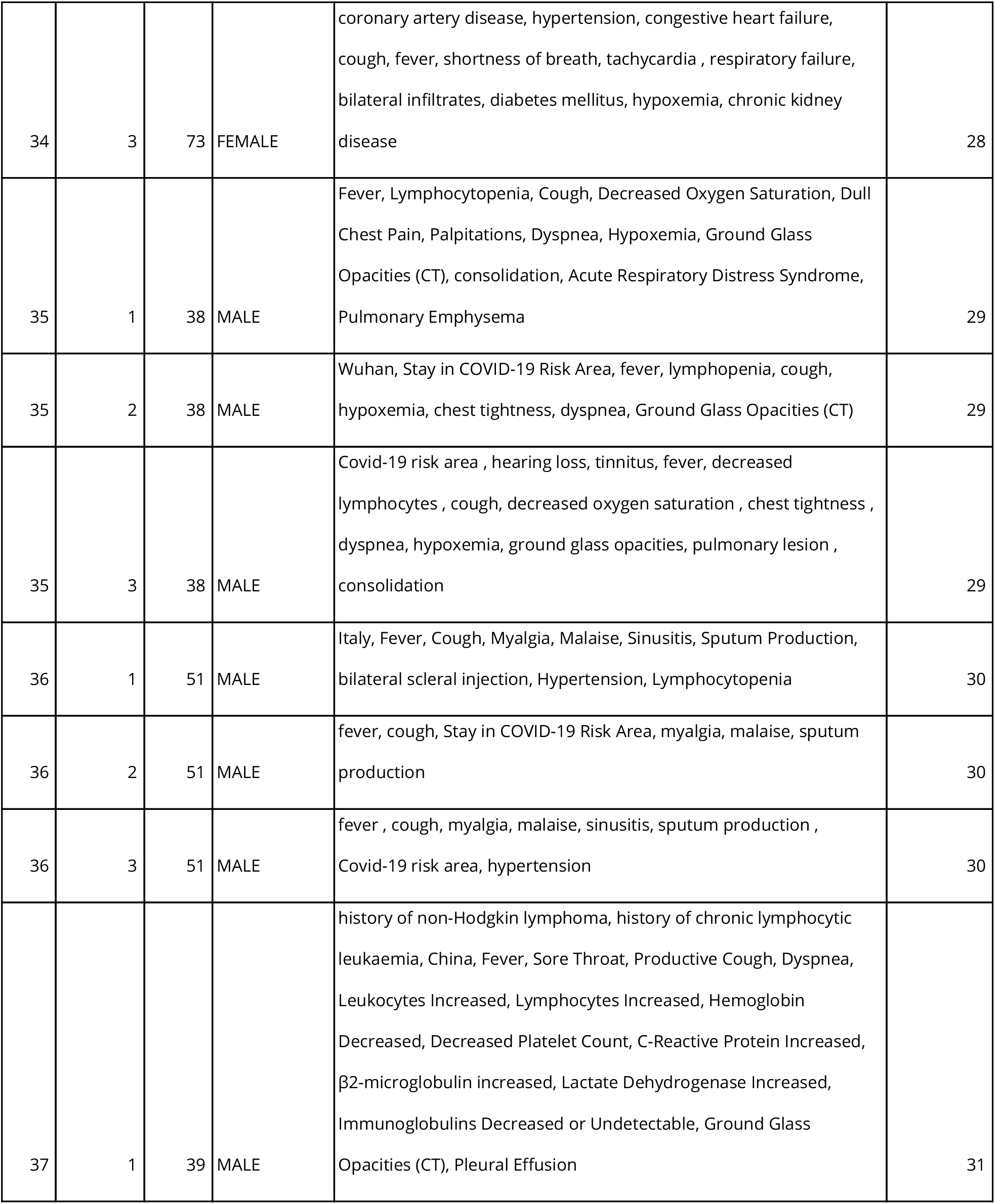
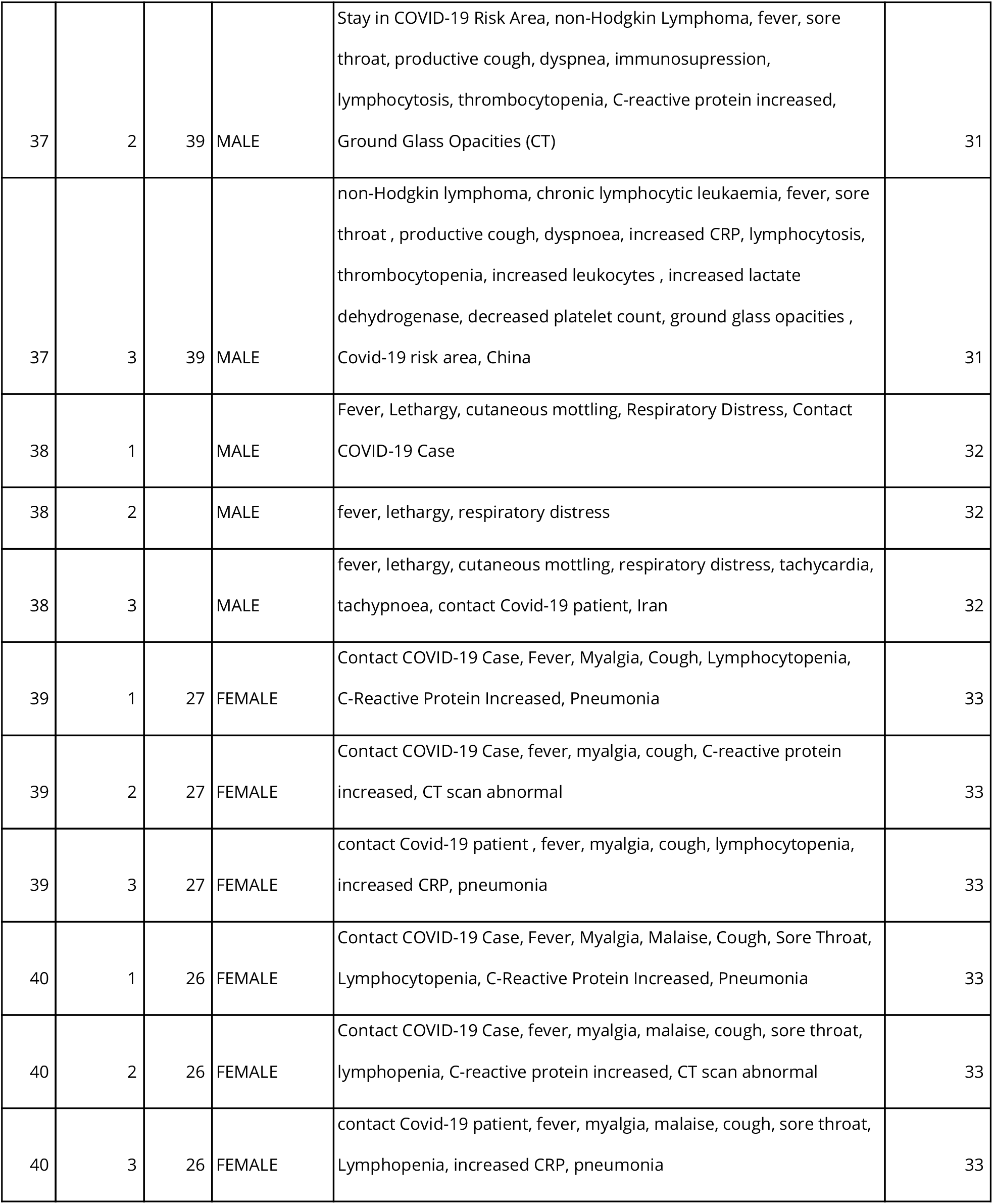
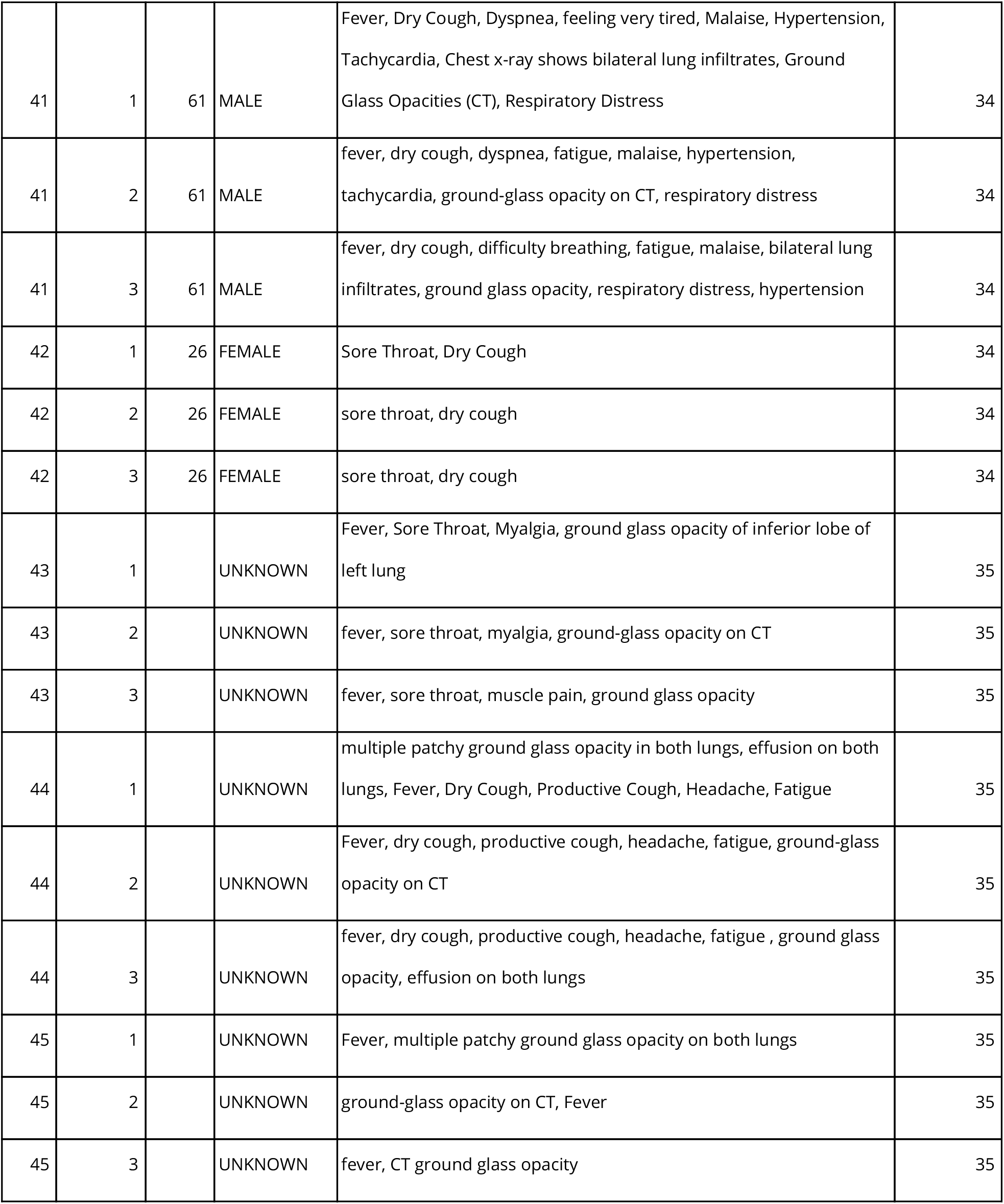
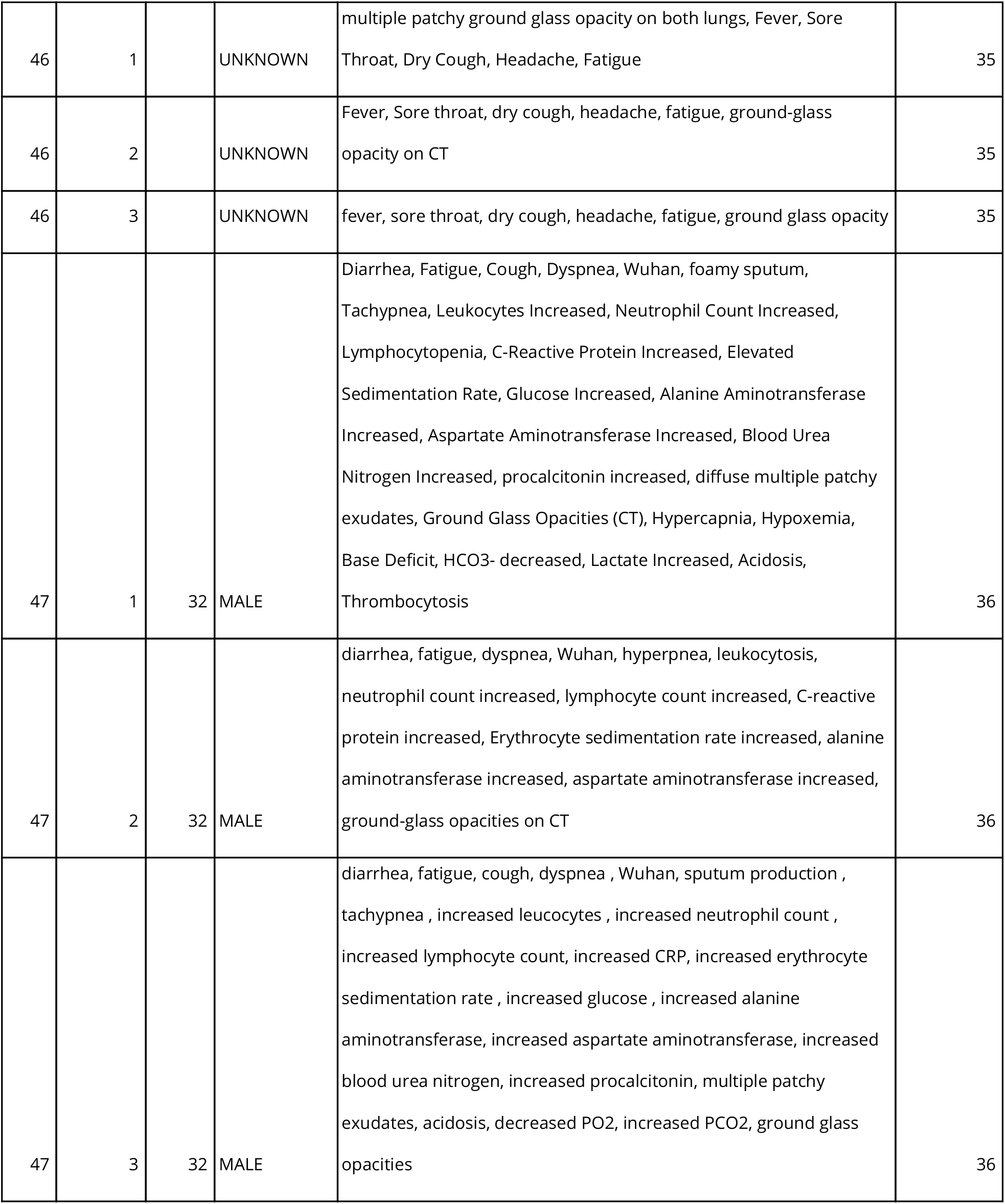
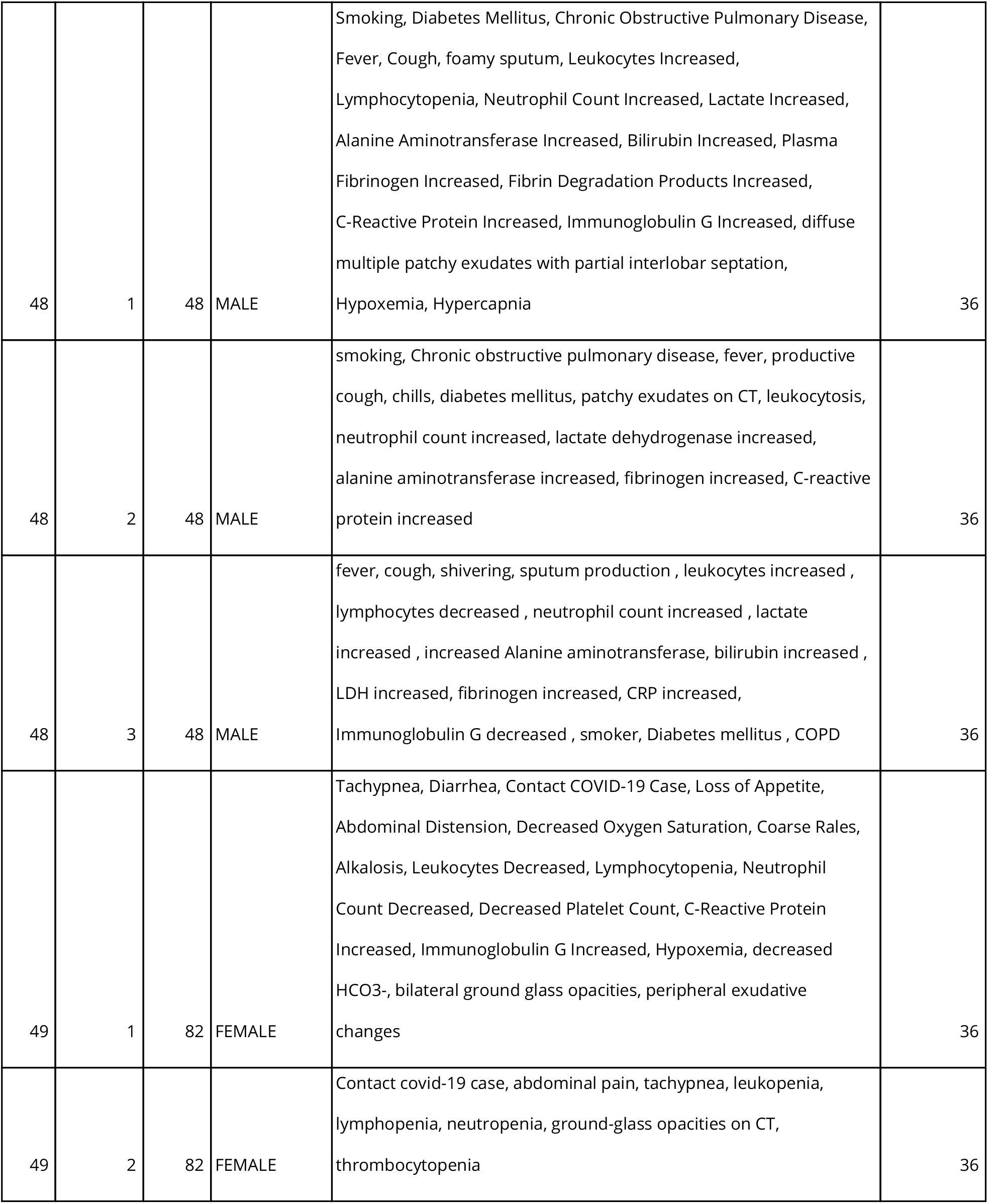
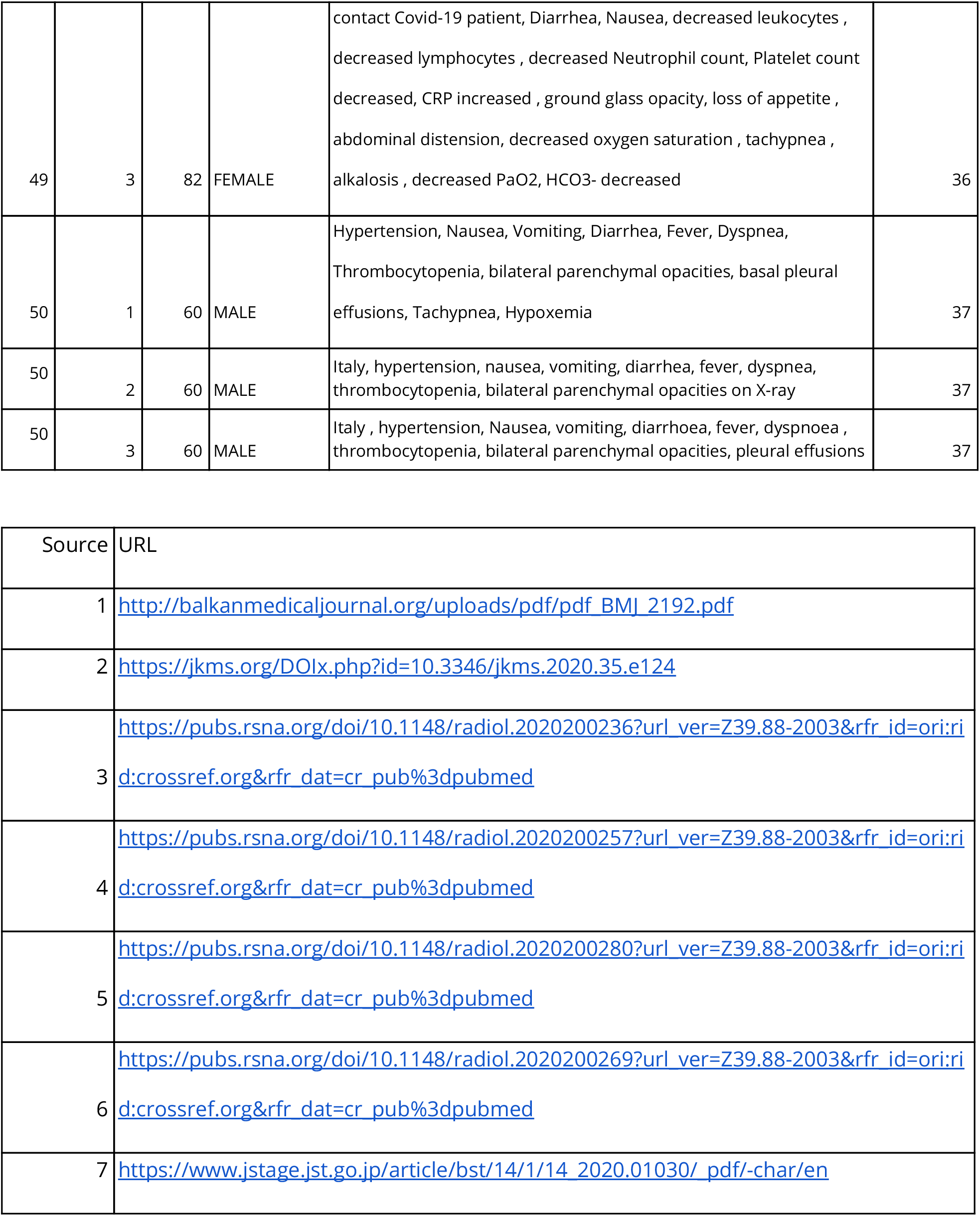
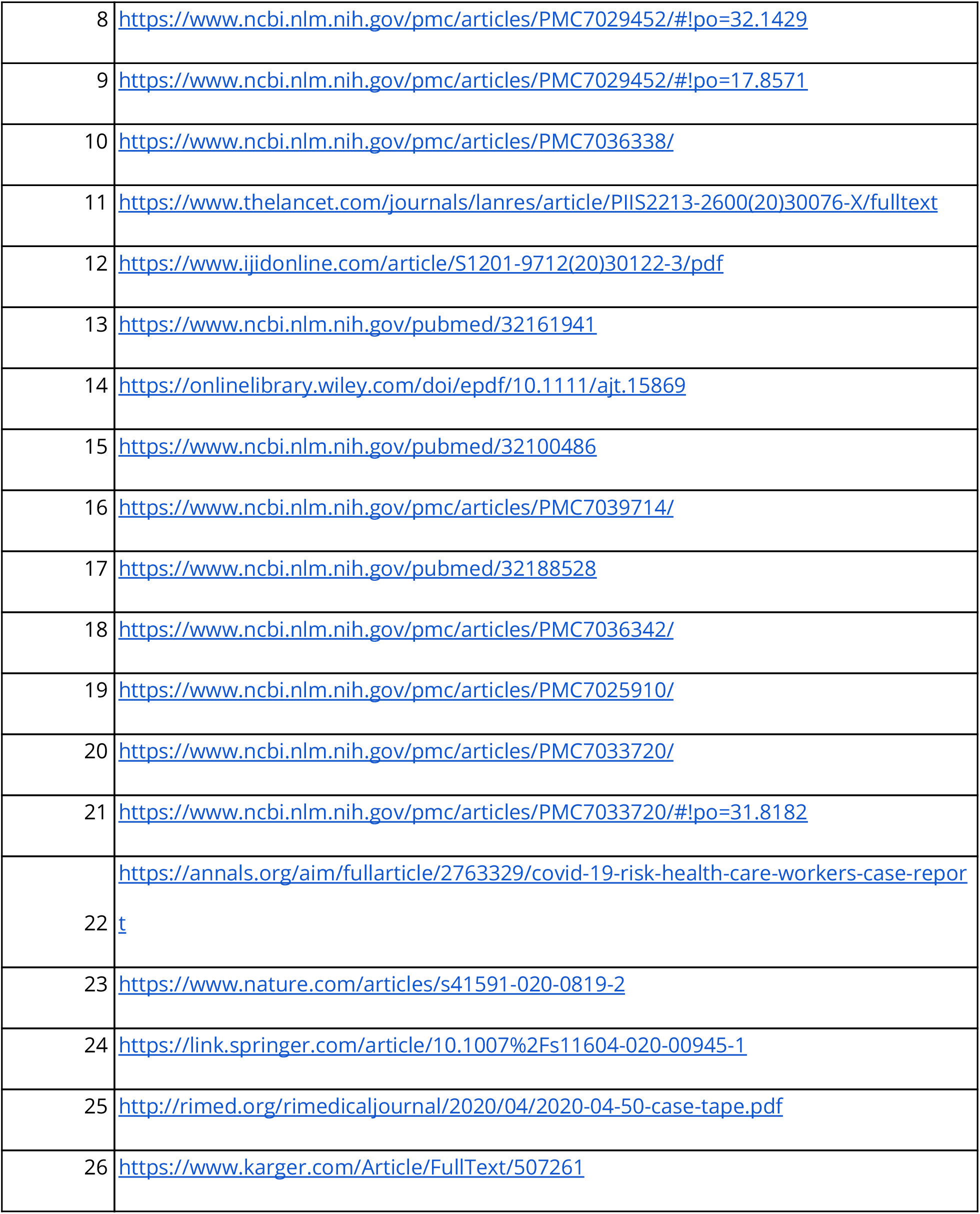
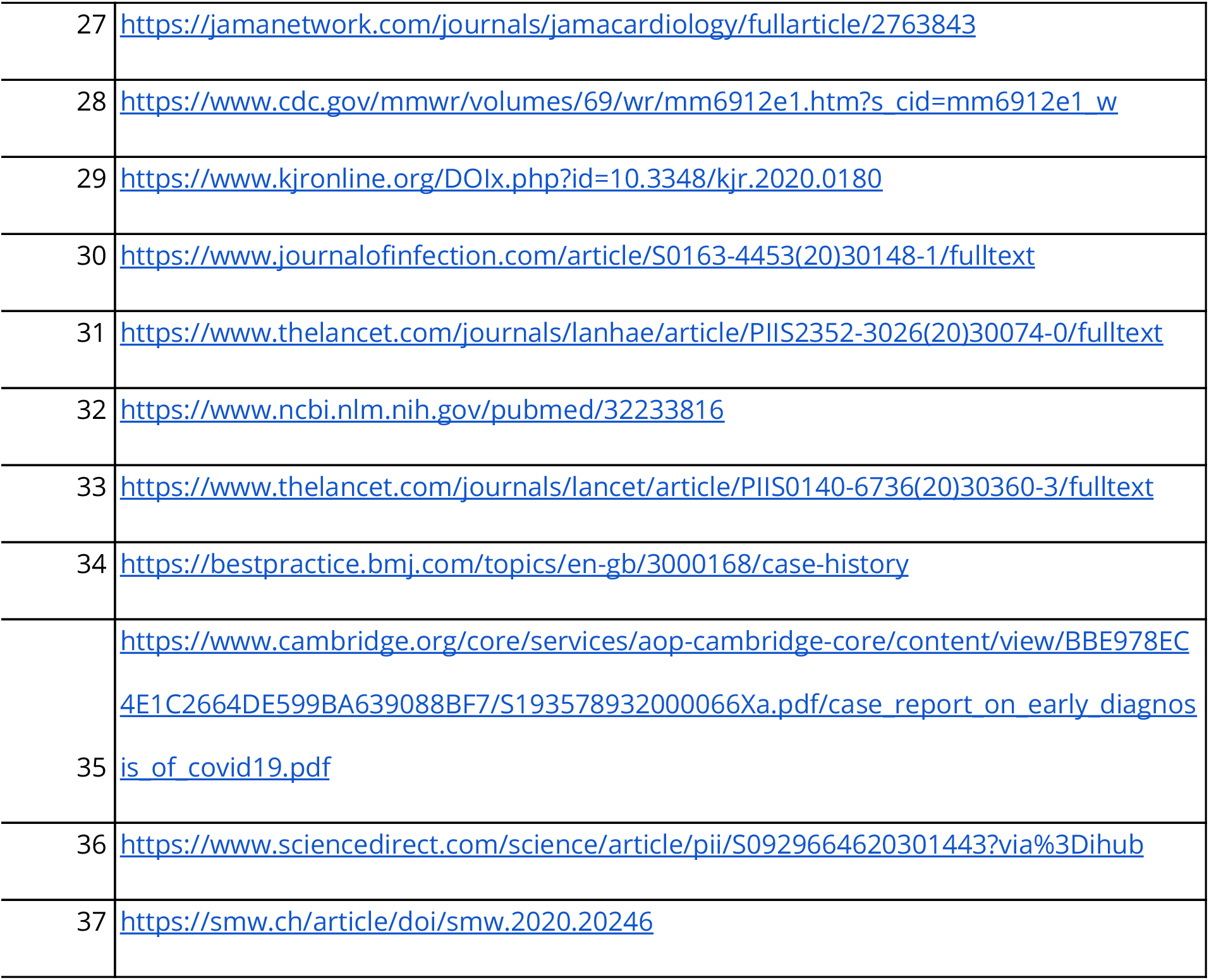
List of the COVID-19 cases

**S4 Table.** List of COVID-19 symptoms according to the WHO

|  |
| --- |
| Fever |
| Fatigue |
| Cough |
| Sneezing |
| Malaise |
| Rhinorrhea |
| Sore throat |
| Diarrhea |
| Headache |
| Dyspnea |

**S5 Table.** Mapping between symptom checkers output texts and risk levels. All mappings were independently done by two different persons and conflicts resolved by a third person’s opinion.

|  |  |  |
| --- | --- | --- |
| Ada | Your symptoms **are probably caused by other diseases** that are more common than COVID-19 | Low |
|  | It's **unlikely** that you're experiencing symptoms of COVID-19 | Low |
|  | Based on what you've reported, you should take steps to monitor your health and practice social distancing. You have **none of the typical symptoms of COVID-19**. However, in the last 14 days, you've been in contact with a confirmed or probable COVID-19 case. Also, you're in one of the groups at risk of developing a more serious form of the disease. | Medium |
|  | The symptoms you have reported are **present in some cases** of COVID-19. You haven't reported exposure to cases of COVID-19. However, you are in one of the groups at risk of developing a more serious form of the disease. | Medium |
|  | You have **symptoms typical of COVID-19**, and you're in one of the groups at risk of having a more serious form of the disease. | Medium |
|  | It's **possible that COVID-19** is causing your symptoms | Medium |
|  | There is a **high risk** that COVID-19 is causing your symptoms | High |
| Apple | You Should Practice Social Distancing | Low |
|  | You Should Self-Isolate | Medium |
|  | Contact Your Healthcare Provider | High |
| Babylon | It's **unlikely** that you are experiencing symptoms of coronavirus (COVID-19). But if you develop a new or continuous cough, fever or difficulty breathing, start a new symptom check. | Low |
|  | The symptoms you mentioned sound worrying and **could be caused** by coronavirus (COVID-19). Call 999 for an ambulance immediately and let them know you may have symptoms of coronavirus. | Medium |
| CDC | Sorry you're feeling ill. Stay at home and monitor your symptoms. Call your provider if you get worse (No COVID-19 risk) | Low |
|  | Stay home and take care of yourself. Call your provider if you get worse. You have one or more symptom that **may be related** to COVID-19 | Medium |
| Cleveland Clinic | You're at **low risk** for COVID-19. | Low |
|  | You're at **medium risk** for COVID-19. | Medium |
|  | You're at **high risk** for COVID-19. | High |
| Docyet | "Es besteht kein Grund zur Sorge! Basierend auf Ihren Angaben ist es **ziemlich unwahrscheinlich**, dass Sie sich mit dem Coronavirus infiziert haben. Sie zeigen keine Symptome von COVID-19. | Low |
|  | Aktuell besteht kein wesentlicher Grund zur Sorge. Basierend auf Ihren Angaben ist es **ziemlich unwahrscheinlich**, dass Sie sich mit dem Coronavirus infiziert haben. | Low |
|  | Aktuell besteht kein wesentlicher Grund zur Sorge. Basierend auf Ihren Angaben ist es **relativ unwahrscheinlich**, dass Sie sich mit dem Coronavirus infiziert haben. Sie schildern jedoch einige typische Symptome eines Atemwegsinfekts (z.B. Grippe, Bronchitis) oder einer Erkältung. | Low |
|  | Sie sollten dringend ärztliche Hilfe aufsuchen. Sie haben typische Symptome einer schweren Grippe oder Lungenentzündung geschildert. Auf Basis Ihrer Angaben ist es jedoch **nicht sehr wahrscheinlich**, dass Sie sich mit dem Coronavirus infiziert haben. | Low |
|  | Sie zeigen deutliche grippeähnliche Symptome und waren potentiell in Kontakt mit am Coronavirus infizierten Personen. Es ist **daher möglich**, dass auch Sie sich angesteckt haben. | Medium |
|  | Sie zeigen erste grippeähnliche Symptome und waren potentiell in Kontakt mit am Coronavirus infizierten Personen. Es ist **daher möglich**, dass auch Sie sich angesteckt haben. | Medium |
|  | "Bisher zeigen Sie keine auffälligen Symptome. Sie sind jedoch in Kontakt mit am Coronavirus infizierten Personen gekommen. Es ist **daher möglich**, dass auch Sie sich angesteckt haben, auf Grund der zweiwöchigen Ausbruchszeit jedoch noch keine Symptome zeigen. | Medium |
|  | Ihre Symptome deuten auf eine schwere Grippe bzw. Lungenentzündung hin. Außerdem waren Sie potentiell in Kontakt mit am Coronavirus infizierten Personen. Es ist **möglich**, dass auch Sie sich angesteckt haben. | Medium |
|  | "Sie zeigen deutliche grippeähnliche Symptome sowie erste Anzeichen einer Lungenentzündung. Außerdem waren Sie potentiell in Kontakt mit am Coronavirus infizierten Personen. Es ist **daher möglich**, dass auch Sie sich angesteckt haben. | Medium |
| Providence | You do not report exposure to coronavirus (COVID-19) and do not have symptoms. You don't report any symptoms of coronavirus infection, so your risk is **low**. You can use this tool again or call your provider if anything changes, but in the meantime: | Low |
|  | You **might be** infected with coronavirus (COVID-19). Please do one of the following: Call 911 for a life-threatening emergency. Schedule an evaluation with your primary care physician. Speak with a provider at Express Care Virtual. | Medium |
|  | Please do one of the following: Call 911 for a life-threatening emergency. Schedule an evaluation with your primary care physician. Speak with a provider at Express Care Virtual. | High |
| Symptoma | **High risk** for COVID-19 | High |
|  | **Medium risk** for COVID-19 | Medium |
|  | **Low risk** for COVID-19 | Low |
| Infermedica | Your symptoms **do not suggest** that you have COVID-19. Continue following the common measures and government directives to avoid contracting COVID-19. Remember that your symptoms may also result from other diseases and may require medical consultation - this interview targets the COVID-19 infection. If your symptoms seem severe and you are worried, contact your doctor or local health authorities. | Low |
|  | Your symptoms are worrisome and **may be related** to COVID-19. Call your local COVID-19-related healthcare number. Depending on the country, this may be a telephone line issued by the Ministry of Health or Health Department. | Medium |
| Your.MD | **There's nothing at present to suggest** that you have coronavirus (COVID-19). Please practice physical/social distancing. | Low |
|  | You **may** have coronavirus (COVID-19). | Medium |
|  | You **may** still be at risk of coronavirus (COVID-19). | Medium |
|  | You're **highly likely** to have coronavirus (COVID-19). | High |
|  | Call an ambulance immediately. Please tell them you have symptoms that may be caused by coronavirus (COVID-19). | High |

**S6 Table.** Full table of sensitivity, specificity, accuracy, F1 score and MCC for all symptom checkers (COVID-19 positive defined by “high risk” for non binary symptom checkers)

| Symptom checker | sensitivity | specificity | accuracy | F1 Score | MCC |
| --- | --- | --- | --- | --- | --- |
| Ada | 0.14 | 1.00 | 0.57 | 0.24 | 0.27 |
| Apple | 0.22 | 0.69 | 0.46 | 0.29 | -0.10 |
| Babylon | 0.90 | 0.33 | 0.62 | 0.70 | 0.29 |
| CDC | 0.94 | 0.29 | 0.61 | 0.71 | 0.30 |
| Cleveland Clinic | 0.32 | 0.74 | 0.53 | 0.40 | 0.07 |
| Docyet | 0.16 | 1.00 | 0.58 | 0.27 | 0.29 |
| Infermedica | 0.78 | 0.83 | 0.80 | 0.80 | 0.61 |
| Providence | 0.32 | 0.72 | 0.52 | 0.40 | 0.05 |
| SF-COS | 0.90 | 0.66 | 0.78 | 0.80 | 0.58 |
| SF-DIST | 0.52 | 0.70 | 0.61 | 0.57 | 0.22 |
| Symptoma | 0.88 | 0.97 | 0.93 | 0.92 | 0.85 |
| Your.MD | 0.14 | 1.00 | 0.57 | 0.24 | 0.27 |

**S7 Table.**
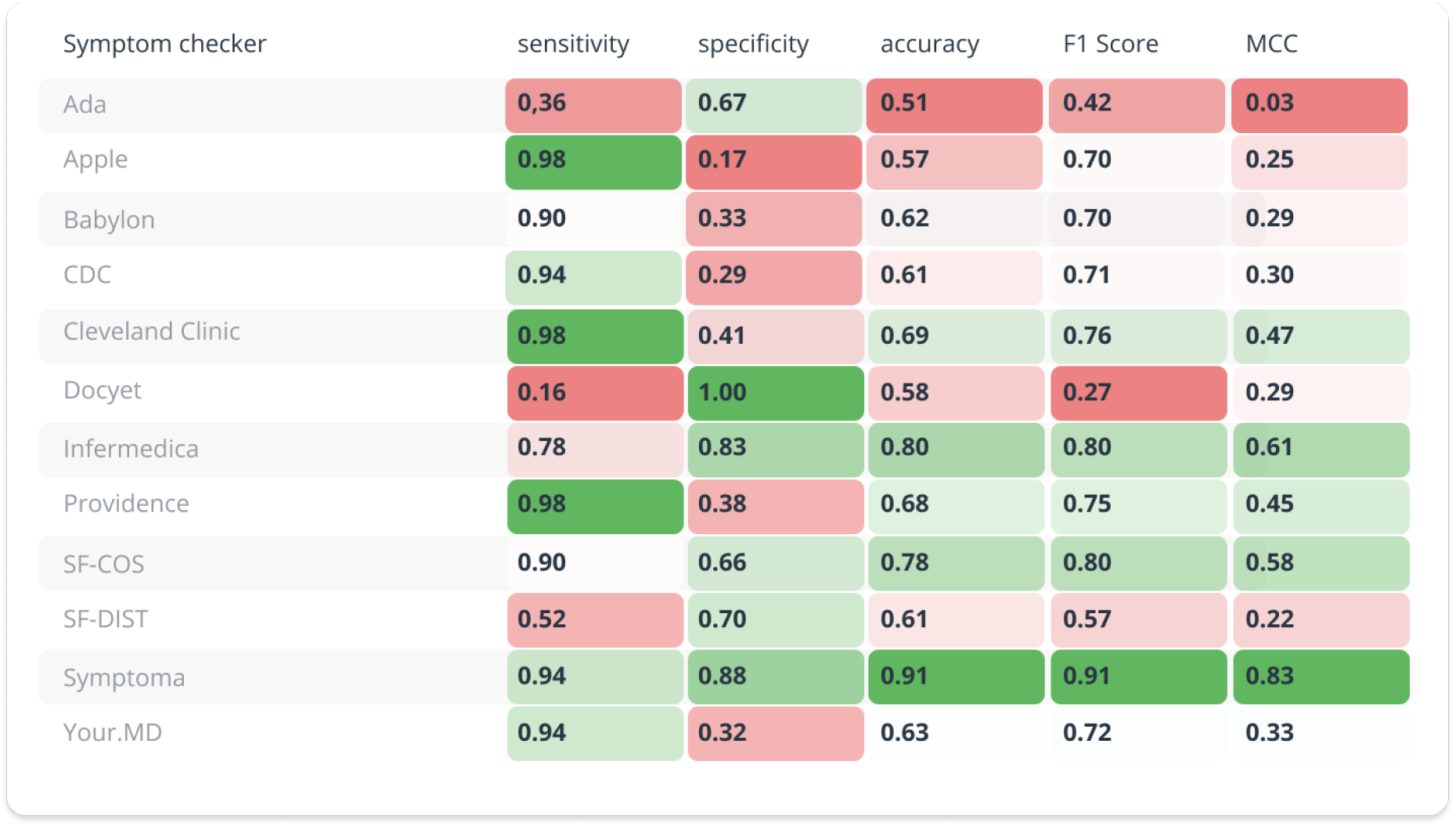
Full table of sensitivity, specificity, accuracy, F1 score and MCC for all symptom checkers (COVID-19 positive defined by “medium risk or “high risk” for non binary symptom checkers)

**S8 Fig.**
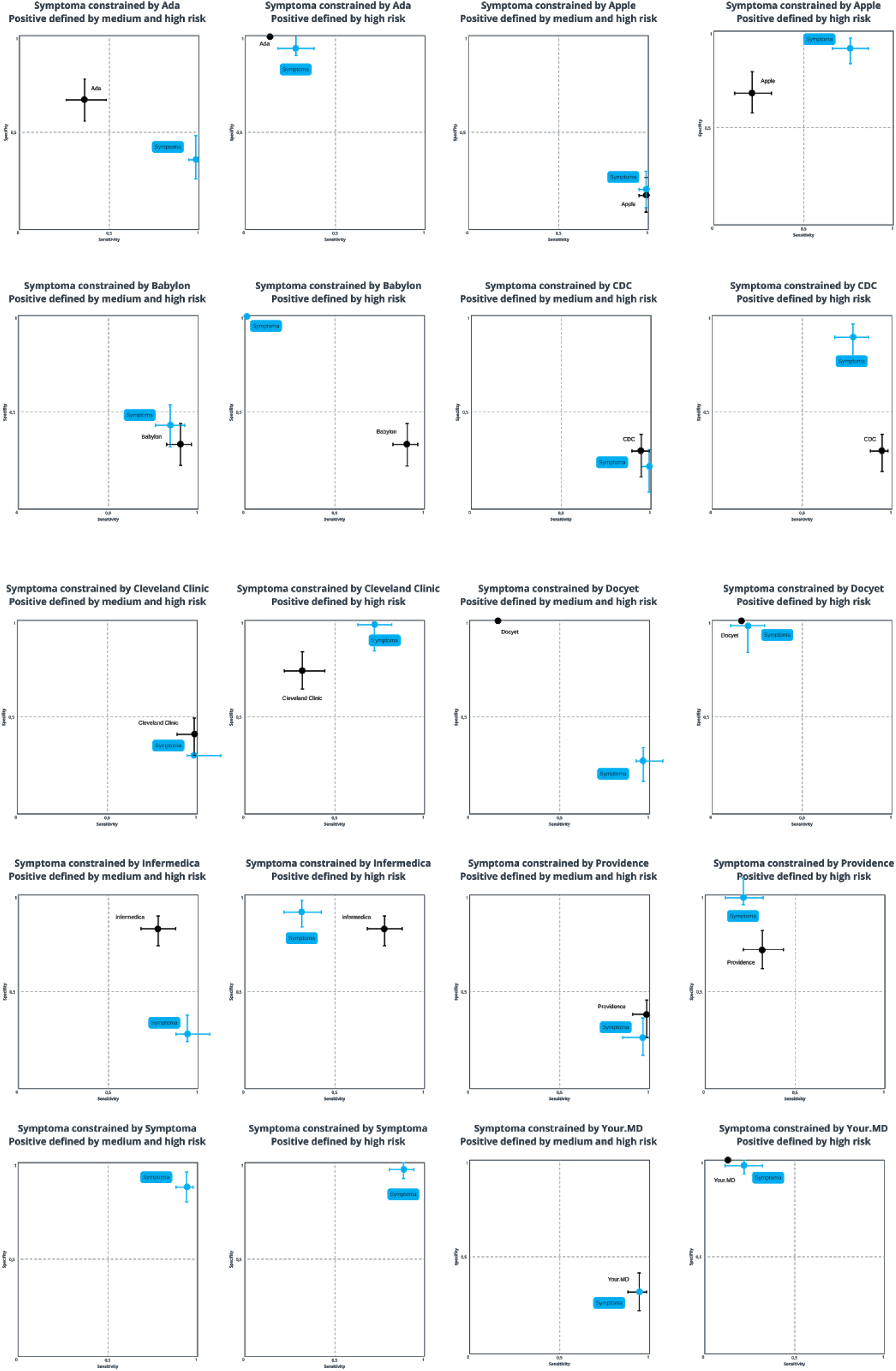
Sensitivity vs specificity for all symptom checkers and Symptoma input constraint respectively by each symptom checker

**S9 Table.** Full table of sensitivity, specificity, accuracy, F1 score and MCC for Symptoma constrained by each symptom checker (COVID-19 positive defined by “high risk” for non binary symptom checkers)

| Symptoma constrained by | sensitivity | specificity | accuracy | F1 Score | MCC |
| --- | --- | --- | --- | --- | --- |
| Ada | **0.28** | **0.94** | **0.61** | **0.42** | **0.30** |
| Apple | **0.76** | **0.92** | **0.84** | **0.82** | **0.69** |
| Babylon | **0.00** | **1.00** | **0.50** | **0.00** | **n/a** |
| CDC | **0.78** | **0.89** | **0.83** | **0.82** | **0.68** |
| Cleveland Clinic | **0.72** | **0.99** | **0.85** | **0.83** | **0.73** |
| Docyet | **0.20** | **0.98** | **0.59** | **0.33** | **0.29** |
| Infermedica | **0.32** | **0.92** | **0.62** | **0.45** | **0.30** |
| Providence | **0.22** | **0.99** | **0.60** | **0.35** | **0.32** |
| Your.MD | **0.22** | **0.98** | **0.60** | **0.35** | **0.31** |

**S10 Table.** Full table of sensitivity, specificity, accuracy, F1 score and MCC for Symptoma constrained by each symptom checker (COVID-19 positive defined by “medium risk” or “high risk” for non binary symptom checkers)

| Symptoma constrained by | sensitivity | specificity | accuracy | F1 Score | MCC |
| --- | --- | --- | --- | --- | --- |
| Ada | **0.98** | **0.36** | **0.67** | **0.75** | **0.44** |
| Apple | **0.98** | **0.20** | **0.59** | **0.70** | **0.28** |
| Babylon | **0.84** | **0.43** | **0.63** | **0.70** | **0.30** |
| CDC | **0.98** | **0.21** | **0.59** | **0.71** | **0.29** |
| Cleveland Clinic | **0.98** | **0.30** | **0.64** | **0.73** | **0.38** |
| Docyet | **0.96** | **0.27** | **0.62** | **0.71** | **0.32** |
| Infermedica | **0.94** | **0.28** | **0.61** | **0.71** | **0.29** |
| Providence | **0.96** | **0.26** | **0.61** | **0.71** | **0.31** |
| Your.MD | **0.94** | **0.32** | **0.63** | **0.72** | **0.33** |

**S11 Fig.**
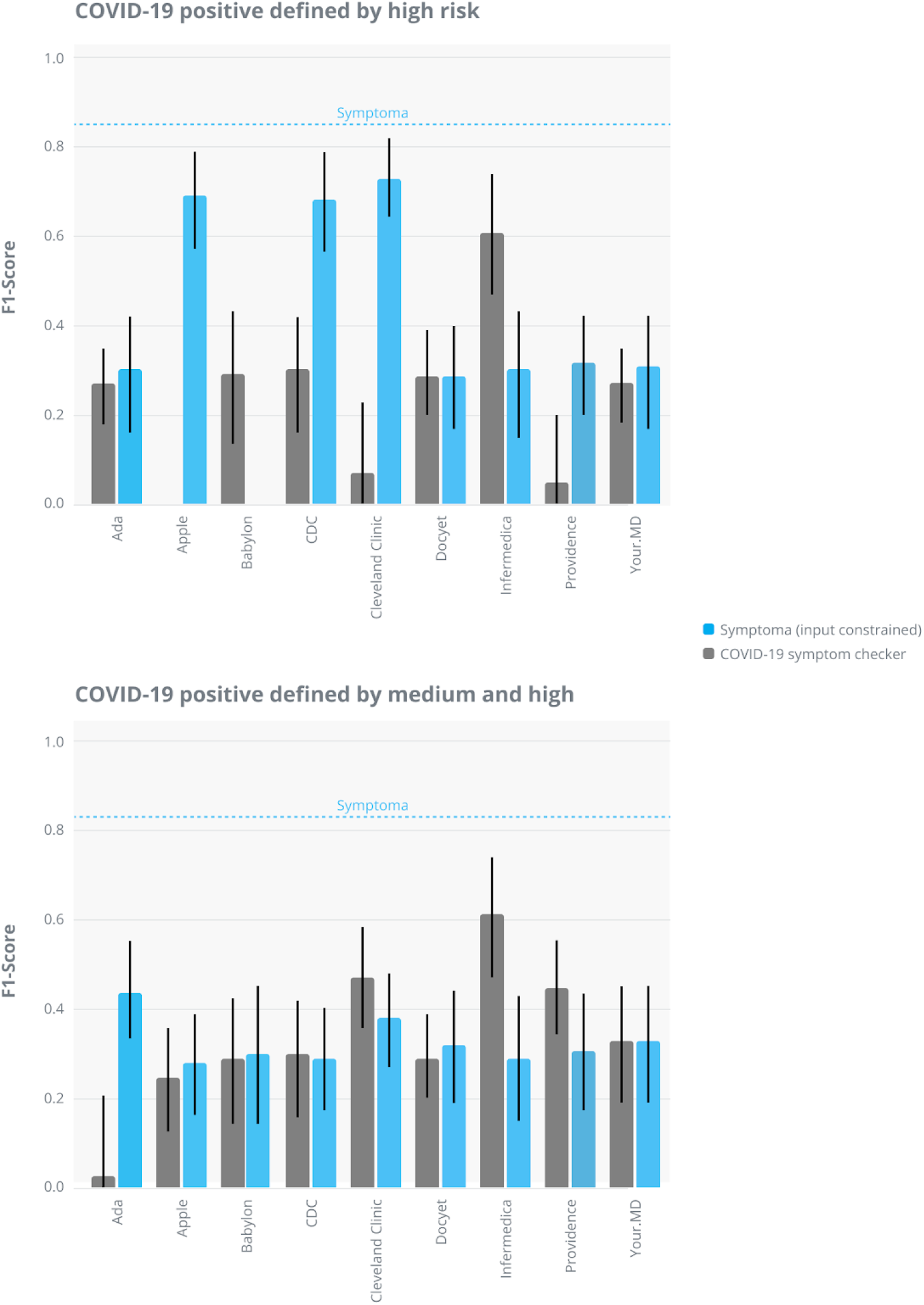
Pairwise comparison between all symptom checkers and Symptoma based on the MCC if only the subset of symptoms used by one checker is also used for Symptoma.

